# A Hierarchical Bayesian Model for Cyber-Human Assessment of Rehabilitation Movement

**DOI:** 10.1101/2022.05.25.22275480

**Authors:** Tamim Ahmed, Thanassis Rikakis, Setor Zilevu, Aisling Kelliher, Kowshik Thopalli, Pavan Turaga, Steven L. Wolf

## Abstract

**Background:** The evidence-based quantification of the relation between changes in movement quality and functionality can assist clinicians in achieving more effective structuring or adaptations of therapy. Facilitating this quantification through computational tools can also result in the generation of large-scale data sets that can inform automated assessment of rehabilitation. Interpretable automated assessment can leave more time for clinicians to focus on treatment and allow for remotely supervised therapy at the home.

**Methods:** In our first experiment, we developed a rating process and accompanying computational tool to assist clinicians in following a standardized movement assessment process relating functionality to movement quality. We conducted three studies with three different versions of the computational rating tool. Clinicians rated task, segment, and movement feature performance for 440 videos in which stroke survivors executed standardized upper extremity therapy tasks related to functional activities. In our second experiment, we used the 440 rated videos, in addition to 140 videos of unimpaired subjects performing the same tasks, to improve our previously developed automated assessment ensemble model that automatically generates segmentation times and task ratings across impaired and unimpaired movement. The automated assessment ensemble integrates expert knowledge constraints into data driven training though a combination of HMM, transformer, MSTCN++, and decision tree computational modules. In our third experiment, we used the therapist and automated ratings to develop a four-layer Hierarchical Bayesian Model (HBM) for computing the statistical relation of movement quality changes to functionality. We first calculated conditional layer probabilities using clinician ratings of task, segment, and movement features. We increased the granularity of observation of the HBM by formulating Δ*_HBM_*, a correlation graph between kinematics and movement composite features. Finally, we used k-means clustering on the Δ*_HBM_* to identify three clusters of features among the 16 movement composite and 20 kinematic features and used the centroid of these clusters as the weights of the input data to our computational assessment ensemble.

**Results:** We evaluated the efficacy of our rating interface in terms of inter-rater reliability (IRR) across tasks, segments, and movement features. The third version of the interface produced an average IRR of 67%, while the time per session (TPS) was the lowest of the three studies. By analyzing the ratings, we were able to identify a small number of movement features that have the highest probability of predicting functional improvement. We evaluated the performance of our automated assessment model using 60% impaired and 40% unimpaired movement data and achieved a frame-wise segmentation accuracy of 87.85±0.58 and a block-segmentation accuracy of 98.46±1.6. We also demonstrated the performance of our proposed HBM in correlation to clinician’s ratings with a correlation over 90%. The HBM also generates a correlation graph, Δ*_HBM_* that relates 16 composite movement features to the 20 kinematic features. We can thus integrate the HBM into the computational assessment ensemble to perform automated and integrated movement quality and functionality assessment that is driven by computationally extracted kinematics.

**Conclusions:** Combining standardized clinician ratings of videos with knowledge based and data driven computational analysis of rehabilitation movement allows the expression of an HBM that increases the observability of the relation of movement quality to functionality and enables the training of computational algorithms for automated assessment of rehabilitation movement. While our work primarily focuses on the upper extremity of stroke survivors, the models can be adopted to many other neurorehabilitation contexts.

## Introduction

Establishing the detailed relationship between changes in function and the underlying changes in movement quality remains a significant challenge in neurorehabilitation [1, 2]. During assessment and training rehabilitation, clinicians are limited in the number of impaired movement elements to which they can attend. Consequently their ability to connect their expert observations to standardized norms or values [3] is limited. Depending on their training and experience, clinicians focus on different movement elements when assessing patients during either direct interactions or when observing videos of patient performance [4, 5]. This level of subjective assessment contributes to the function/quality challenge in the absence of a normative quantitative framework [6, 7].

The evidence-based quantification of the relation between changes in movement and function can assist clinicians in achieving more effective structuring or adaptations of therapy [1]. This assistance could result in the generation of large scale data sets to automate assessment, thus leaving more time for clinicians to focus on treatment and allowing for remotely supervised therapy at the home [8]. This quantification can be partially achieved through the detailed tracking of kinematics and their correlation to validated, expert driven, clinical measures [2, 9]. Although high-end sensing technologies can provide the necessary detailed tracking, these technologies are cumbersome even in the clinic, and certainly not yet feasible for the home. Tracking of movement through marker-based capture or intricate exoskeletons is costly, complex, and obtrusive [10, 11, 12].

The home environments of many stroke survivors are also often constrained and cannot easily accommodate the type of large-form technology typically used in hospital or clinic rehabilitation systems [13]. Even small systems, if intrusive or perceived negatively, tend to be rejected by the stroke survivor and/or their care partner [13]. Therefore, kinematic data need to be captured through low-cost and unobtrusive means if high-fidelity data are to be acquired. Currently, the effort needed to capture patient data, combined with variations in the quality of patient movement, presents profound limitations in the utility of such data.

Our team proposes a Hierarchical Bayesian Model (HBM) for computing the relationship of movement quality to functionality in rehabilitation. The proposed HBM leverages expert clinician knowledge to constrain the computational challenge of working with limited, low fidelity, and high variability data. The model also leverages computation to assist the clinician in integrated observation of function and movement quality. In the following sections we discuss the development of the model in detail. We show how the model can help establish evidence-based quantifiable relations between movement changes and function and how these relationships can then be used for the automated assessment of movement. Summaries of the assessment are subsequently provided to the clinician to assist in structuring and adapting therapy in the clinic, and remotely, in the home. We focus our presentation on the application of the model to the rehabilitation of the upper extremity of stroke survivors. However, the model can be adapted to many other neurorehabilitation contexts.

## Making the clinician assessment process more observable

Technological systems aiming to assist the clinician in delivering rehabilitation may not be adopted by clinicians if they are incompatible with the clinicians’ approaches or introduce steep learning curves [14]. Busy clinicians have limited training time to learn, troubleshoot, and maintain complex new systems. Over the past few years, our team has used participatory design processes [15] and custom-made interactive video rating tools [16] to help expert clinicians reflect on and reveal their movement assessment processes and the related internalized (tacit) rating schema so that we can base our cyber-human assessment models on observable expert schema. Our work reveals that clinicians use a hierarchical probabilistic process for dealing with the uncertainty and complexity of therapy assessment.

Figure 1 presents a representational approximation of the clinician observation and decision processes. At the highest layer of the hierarchy, we place the movement impairment level of the patient. The level of impairment is intrinsically on a continuum; for convenience it is organized into three base categories – mild, moderate, severe – and category combinations – mild/moderate and moderate/severe – allowing for a total of five categories. This categorization is based on validated clinical tests that use measurements of the components of the physical apparatus (e.g. Fugl-Meyer test) [17], observations of timed-tasks in the clinic (e.g. Action Research Arm Test (ARAT) [18]), and Activities of Daily Living (ADL) questionnaires (e.g. Motor Activity Log [19]). The therapy is effective when the impairment assessments based on ADL instruments, and the impairment assessment in the clinic, are both reduced in a highly correlated manner (the more effective the therapy, the greater the impact it has on daily living).

**Figure 1:** (Available per request) Different layers of the Hierarchical Bayesian Model (HBM). The functionality decreases as we go down the hierarchy and movement quality decreases as we go up. The area where both are greyed out is the most important part of the HBM.

However, detailed and accurate observations (and potential quantification) of ADLs are challenging. The clinician needs to rely on patient questionnaires, which in turn rely on patient memory and perception, and can thus be subjective and imprecise [1, 20]. To help increase the impact of therapy on daily functionality, clinicians utilize sets of generalizable therapy tasks that map well to ADLs. If the performance of these tasks improves during therapy training, then performance of ADLs should also improve, indicating that the impairment level is decreasing. Understanding how clinicians utilize these tasks to address functionality and movement quality is critical in making their assessment processes more observable. We collaborated with expert clinicians to define 15 generalizable training tasks for upper extremity rehabilitation, ranging from simple reach-to-touch tasks to more complex transportation and bimanual manipulation tasks. We also developed a rating rubric for these tasks. The rubric follows well established paradigms (e.g., WMFT, ARAT etc.) with task ratings ranging from 0 to 3, where 0 denotes that the task was not attempted and 3 indicates a close to unimpaired performance. We place the generalizable training tasks at the second layer of our hierarchy. Performance on a task is characterized by a label (*T*_2_) (i.e. Task 2 of the 15 training tasks) and the clinician Rating (*T*_2_*_,r_*) for that performance (i.e. a rating of 0 to 3). We use a dotted line representation of the relation of the observed tasks to ADLs to denote that the relation is not fully observable.

We place detailed raw kinematics of movement during therapy (which are captured with low cost/low intrusion technology) at the bottom of our hierarchy and represent it as a two-dimensional matrix. The y-axis represents the different patients and the x-axis the multitude of movement features that need to be tracked. Following related work by [21], we denote the representation of this lowest level as data (D). Looking at the top and bottom of our hierarchy, any one indication of impairment (i.e. some level of moderate impairment) is probably due to a high number of different combinations of low level features that cannot be fully observed in real time by a clinician [6]. At best, a clinician can focus on a few movement elements and describe them as impaired, mildly impaired or not impaired, without quantifying this categorization in detail. In our matrix visualization, we show this coarse categorization using a grayscale schematic where black is impaired, grey is mildly impaired, and white is not impaired.

To manage the complexity of real time movement observation and to make generalizable observations across different therapy tasks, clinicians tend to segment tasks into a small set of generalizable segments that can be combined through different paths to generate the therapy tasks. Even though most clinicians use intuitive segmentation of movement for observation and assessment, the segment vocabulary is not standardized. We worked with expert clinicians to create a standardized segment vocabulary that can produce the 15 training tasks used in our model as well as many other upper extremity therapy tasks that map to ADLs. The segments are: Initiation + Progression + Termination (IPT), Manipulate & Transport (M&TR), Complex Manipulation and Transport (CMTR), and Release and Return (R&R). As an example, a drinking related task can be described by the following codification: subject reaches out and grasps a cone object (IPT) and brings it to their mouth (M&TR), then returns the object to the original position (M&TR), and releases the object and returns the hand to the rest position (R&R).

For computational purposes, the devised segment vocabulary and the combination paths that allow the compilation of the tasks can be represented as a simple state machine (see Figure 2). To make assessment of segments in real-time, the clinician significantly limits the features observed per type of segment. This limitation is achieved by using their own experience to develop a probabilistic filtering of irrelevant low-level features for a segment (i.e. digit positioning is likely not that relevant to movement initiation), and a probabilistic composite observation of relevant individual features (i.e. a strategy for quick impressions of shoulder and torso compensation during movement initiation). This process is not well standardized as the filtering and compositing activities are based on individual experience and training. We again worked with expert clinicians to define a consensus-limited set of composite movement features that are important when assessing the performance of each segment in our model. In [16] (Table 3), data from our previous study shows the key composite features per type of segment. For example, the resulting rubric identifies four key features to assess during the Complex Manipulation and Transport stage: i) appropriate initial finger positioning, ii) appropriate finger motion after positioning, iii) appropriate limb motion following finger positioning, and iv) limb trajectory with appropriate accuracy. We also worked with the clinicians to establish operational definitions of the terms used to evaluate the composite movement features. For example, the word “appropriate” used in the above instruction is defined as “the range, direction, and timing of the movement component for the task compared to that expected for the less impaired upper extremity.”

**Figure 2:**
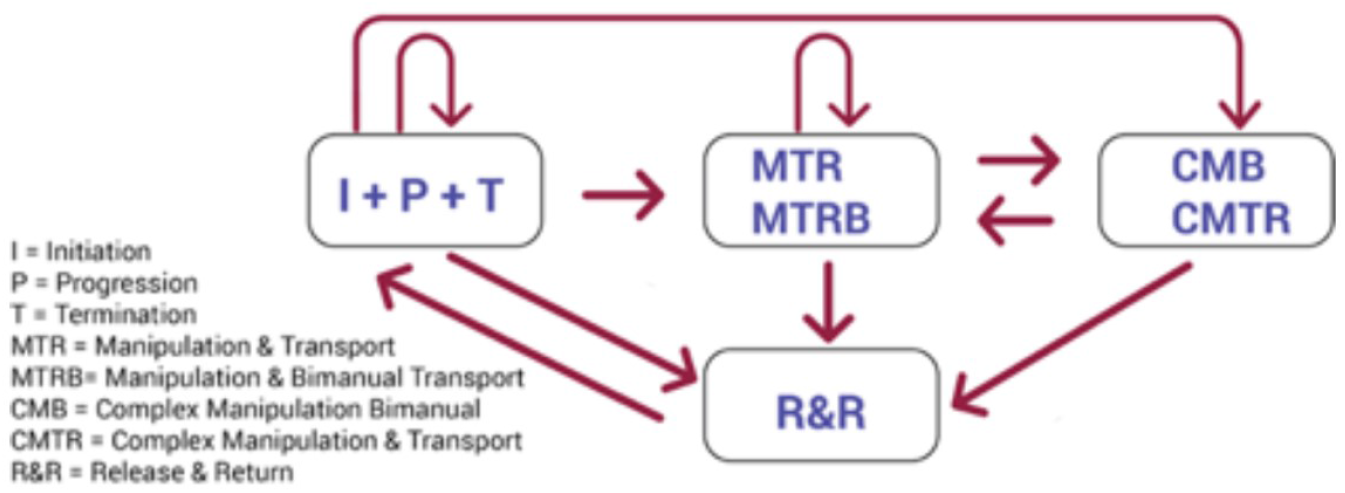
State -achine used to design different activities in the SARAH system.

We can now see how the full hierarchy can be used to begin to reveal the integrative relations between functionality and movement quality. The movement impairment level of the patient depends on the assessment of performance of therapy tasks that map to Activities of Daily Living. The performance of the therapy tasks depends on the performance of different series of standardized segments. The performance of each segment depends on the quality of a small number of composite movement features. The quality of the composite movement features depend on the quality of individual kinematic features. Each layer of the hierarchy conditions the layer below and through the full series, overall functionality during daily life can be connected to the quality of individual kinematic features.

Although our work with clinicians reveals a hierarchical approach to assessing the performance of therapy tasks and establishes the layers of such a hierarchy, the exact relationships of the layers can not yet be established. For example, what is the exact level of torso compensation during initiation of a reach, grasp, and transport task that could impair performance of the initiation segment (and consequently of the overall task) and how does the level of compensation relate to the amount of torso rotation or leaning? Without these relations, a standardized approach to computationally assisted analysis of the connections of function and movement can not be realized.

We also note that clinicians can have a much more informed discussion on moment quality and functionality when reacting to video recordings of movements of patients. Further advancement of modeling requires the collection of video recordings of patient movement and the structured rating of those recordings by clinicians. These ratings could inform the further modeling and standardization of the clinician assessment process. The ratings and the resulting models could then be used to train computational algorithms for the automated assessment of movement quality. We could also formulate the proposed hierarchical Bayesian model to quantify the relation between movement quality and function using the clinician’s ratings and raw kinematics.

## Materials and Methods

Our methodology integrates three processes and three related experiments. In experiment 1, we describe the development and testing of a rating process and tool that helps clinicians standardize a movement assessment approach relating functionality to movement quality. In experiment 2, we describe our machine learning ensemble methodology for the automated segmentation and assessment of patient movement. The automated assessment methodology leverages clinician ratings and the expert knowledge integrated in the ratings to overcome the challenge of working with variable, noisy, and limited data. In experiment 3, we describe a method that combines the clinician ratings with computational analysis to express an HBM that quantifies (in a statistical manner) the relationship of function to movement quality.

### Impaired activity space variant data collection

We collected videos of nine stroke survivors performing 12 of the 15 upper extremity generalizable training tasks that we established through our pilot research. Seven men and two women participated in our study. Two participants were categorized as moderate impairment (Fugl-Meyer score between 30 - 55), while seven participants presented with mild to moderate impairment (Fugl-Meyer score of greater than 55). The nine participants also had different specific movement challenges thus providing an even more varied data set. Each patient was asked to attempt each of the 12 tasks four times. The more severely impaired patients could not complete four iterations. The majority of patients could not perform the more challenging tasks (tasks 11 and 12). The patients were not asked to attempt the most challenging tasks (tasks 13-15). Three of the nine patients were asked to return for two repeat sessions to allow the clinician to explore differences within data sets of the same patients. In total, our 15 data capture sessions produced 600 usable video recordings of single tasks. The movement of the patients was recorded using one low-cost video camera. No detailed instructions were given to the clinicians for setting up the camera. Clinicians were told to set up the camera on the impaired limb side, approximately three feet away. These instructions facilitated an easy setup of the camera without interfering with the therapy sessions. However, this resulted in variant points of view for the camera and a noisy dataset. Such datasets creates significant challenges in training machine learning models and thus require additional pre-processing (as discussed in [22]).

### Unimpaired activity space invariant data collection

To establish a ground truth, we also collected videos of seven unimpaired patients performing the 15 upper extremity generalizable training tasks. Three men and four women participated. All the participants were right limb dominant. Each was asked to attempt each of the 15 tasks two times each. In total, our 15 data capture sessions produced 210 usable video recordings of single tasks. As we were working with unimpaired individuals, we could have a lengthier preparation and system setup time and thus secure better data. To standardize the capture setup and ensure high-quality video capture with activity space invariance, we introduced a real-time activity space calibration step. The details of this setup will be elaborated in a future publication. We also added an additional camera to the capture system. We used both a right sagittal and a frontal camera for the unimpaired data capture as changing the viewing angle to better understand hand-object interaction over time is important. The setup used to capture the unimpaired patient data is shown in [22] (Figure 1). As we are in the initial development stage, all the analysis in this paper were performed using the right sagittal captures.

## Experiment 1: Rating Interface

### Design of the Experiments

The detailed development of the rating process and interactive rating interfaces are described in previous publications [16, 15, 22]. Here, we summarize only the development components that proved critical to the expression of the functionality-movement quality HBM. We developed a custom rating interface, called the Video Application Tool (VAT), using an HTML, CSS, and JavaScript front-end. We stored the patient information on our secure MongoDB server, and encrypted the videos of patients before saving them to individual computers for each clinician. The VAT presents the clinician with the instruction video of an unimpaired person completing each task (which was shown to the patient before attempting each task), and the sagittal view of the stroke survivor/patient then attempting that particular training task.

The VAT allows clinicians to assess the video recordings of tasks through integrative ratings of the three layers of our assessment hierarchy that are readily observable by experts: task performance, segment performance, and composite feature performance per segment. To facilitate effective ratings by clinicians, the composite feature ratings are binary (impaired, not impaired) and the rating of segments and tasks is realized on a 4-point scale similar to scales used for other validated assessment instruments (i.e ARAT, WMFT):

- 0 segment/task not attempted
- 1 incomplete or inaccurate performance of segment/task and/or performance through significant compensation
- 2 complete performance but with noticeable movement impairment or very slow performance
- 3 segment/task performed with no significant movement impairment and within reasonable amount of time.

Through a pilot session with expert clinicians, we established that a skilled clinician could complete one full task rating (overall movement and all movement segments) in five minutes or less. The participating clinicians also concluded that a rater would most likely reliably rate no more than 12 tasks in one sitting/session (owing to possible fatigue or focus issues over time). Thus, the interface allows the clinician to move through 12 full task ratings at a time per session, although they have the option of beginning a new 12 video session upon completion if they wish.

We designed three rating experiments to help the clinicians explore and define the relations of the three layers of our hierarchy and concurrently reveal their tacit understanding of the relationship of functionality (as captured through the rating of the performance of the overall task) with movement quality (as captured through the rating of segments and their related composite movement features). We assessed the three experiments through an interactive participatory design approach. We combined qualitative (semi-structured interviews and longitudinal diary studies) with quantitative statistical analysis to track the relations and potential causation between these three layers. The quantitative analysis focused on: inter-rater reliability (IRR) – the task, segment, and composite feature agreement among clinicians. We calculated IRR using Cohen’s kappa with two raters. Time per session (TPS) - the average time (in minutes) needed to complete the rating of all the videos per session. Score distribution: the distribution of scores among all ratings in each experiment. The three therapist raters had 40, 15, and 12 years of rehabilitation experience each respectively. One rater had assisted with the design and development of the rating rubric, while the other two received in-person and online instructional training to familiarize them with the rubric, the rating schema, and the rating tool. Two therapists participated in the first two experiments, while all three participated in experiment 1c. The third therapist joined the rating team as they will take the lead in co-developing the rating rubric schema for a different future and related upper extremity measurement instrument for the clinic.

#### Experiment 1a: Function to Movement Quality (F2MQ)

Experiment 1a presents our first attempt at understanding how clinicians approach the relation of function to movement quality in rehabilitation. We customized the interface for this experiment though a co-design process with five expert clinicians. All clinicians proposed initially that a function to movement quality rating sequence would best resemble their assessment framework. The score of a task denotes the overall assessment of functionality, while the movement features denoted as impaired by the clinician for each segment provide a more specific interpretive rationale detailing how and why the clinician arrived at their overall score. Thus, the VAT first presents the rater with videos depicting the entirety of the patient performing the task. Next, the rater must assign a discrete score for each movement segment (e.g., Initiation, Progression, and Termination) and selected a checkbox for any of the key movement features of each segment that they assess as impaired. The clinician moves in a linear progression scoring each of the segments. They cannot move from a segment until the rating for that segment is complete. Once they finish rating each of the segments, they are once again presented with the videos depicting the entirety of the patient performing the task, which they must rate again. This feature was implemented at the request of the rubric and rating assessment development team as they were interested in discovering if the process of reflecting on and assessing the movement in discrete segments and individual movement features might compel the clinician to alter their overall score from their initial first assessment. Two clinicians rated 72 tasks in experiment 1a.

The VAT “F2MQ” model interface is displayed in Fig 3 and depicts the interface that the clinician is presented with when viewing the overall training task video. They can choose to view the instructional video or the patient video in the large video panel. The top of the screen presents the linear series of tabs that the clinician selects to move from the overall/total activity video through the segments comprising the complete activity before returning to the overall/total activity video again. When rating tasks, clinicians select one of the rating buttons to provide an overall score. When rating segments, the clinician selects one of the rating buttons and then denotes composite movement features as impaired (using the checkbox next to each feature) to provide interpretive (movement quality) context for the segment rating. In addition, the clinicians can use the “comment” button beneath the main video panel to further annotate their assessment with commentary about the specific video they are viewing. Similarly, the clinicians can use the “flag” button beside it to send a message to the tool development team regarding a technical problem with the interface or an issue with the displayed video. Finally, the progress bar at the bottom of the screen displays where the rating clinician is within a typical rating “session.”

**Figure 3:** (Available per request)VAT (F2MQ model) function to movement quality interface. (a) Overall video assigned rating of 1, with interpretive feature “Task complete, but 2 or more movement elements showed significant impairment; (b) Initiation segment video assigned rating of 1 with interpretive features “shoulder elevation” and “shoulder flexion or abduction” selected, while Progression is rated 2 with interpretive feature “Trunk sway, flexion and rotation” selected. Termination segment is rated 3 with no features selected.

#### Experiment 1b Movement Quality to Function (MQ2F)

The second iteration of the rating process and tool begins to challenge the clinician’s methods of assessing rehabilitation movement while exploring further the conditioning effect of movement quality observations on functionality assessment (Figure 4). In experiment 1b, the overall task and segment scores are computationally generated based on the composite movement feature impairment observations of the clinician. We customized the interface for this experiment though a co-design process with three clinicians. We moved the main video interface panel to the left of the screen with the video selection buttons placed above. This change provided additional screen real-estate to the right of the interface where we could more carefully and prominently display the composite movement features for clinicians to consider. When assessing a segment, a clinician is first asked to check the composite movement features showing a level of impairment that can influence function/task execution. If only one feature is checked as impaired, then a segment score of 2 is automatically generated. If two or more features are checked as impaired, then a segment score of 1 is automatically generated. The clinician can influence the segment score by denoting fewer or more composite movement features as impaired. With input from the three clinicians participating in the co-design process, we instructed each rating therapist as to how to understand the relationship between the meaning of each movement feature and its connection to the score generated by the VAT (MQ2F).

**Figure 4:** (Available per request)VAT (MQ2F model) movement quality to function interface. (a) Overall video assigned rating of 1, with interpretive feature “Task complete, but 2 or more movement elements showed significant impairment; (b) Initiation segment video assigned rating of 1 with interpretive features “shoulder elevation” and “shoulder flexion or abduction” selected, while Progression is rated 2 with interpretive feature “Trunk sway, flexion and rotation” selected. Termination segment is rated 3 with no features selected.

Figure 4(b) displays the interface presented to clinicians when asked to rate the Release and Return movement task segment. After rating all segments of a task, the clinician moves on to rate the overall patient performance of the task. Instead of giving a score from 0-3 for the task, the clinicians selects one of six possible interpretations of overall task performance:

- task not attempted
- task not performed fully
- task complete but two or more movement elements showed significant impairment
- task complete but one movement element showed significant impairment
- task complete but length of execution is long
- task completed without impairment

Depending on which phrase is chosen, the computer automatically generates the score shown in parenthesis next to the phrase. These six phrases were co-designed with expert clinicians so as to facilitate connecting their functionality assessment, denoted through the assessment of task performance, to movement quality issues identified at the segment level. In the situation depicted in Figure 4(a), the clinician has selected “Task complete, but one movement element showed significant impairment,” which generates a score of 2. The feature “Task complete, but the length of execution is long” is also displayed in bold text in this situation as it is the one other option that could also generate a score of 2. If the clinician agrees with the score generated based on their interpretive assessment, they can select the confirm button and move to the next segment. This version of the interface introduces the possibility of a performance score of 0. We had overlooked this possibility in our previous version of the rubric/assessment and rating/annotation tool combined. During the presentation session with the clinical team at the end of Experiment 1a, it became clear that performance events were not well described using our 1 - 3 rating schema, and non-performance needed to be considered. Two clinicians rated 185 tasks for Experiment 1b.

#### Experiment 1c Structured Decision Process (SDP)

In our final iteration, the VAT guides the rating clinician through a structured decision tree process. The binary decision points and their sequence were again co-designed with expert clinicians and informed by validated clinical measures for the standardized assessment of upper extremity therapy of stroke survivors [23]. The process aims to reveal underlying assumptions made by clinicians when assessing movement and assist the rating clinicians in explicitly connecting assessment of functionality and movement quality. Additionally, in this iteration, we wanted to analyze if the process of using the VAT helped each clinician reflect on their practice beyond the use of the VAT.

Clinicians were asked to answer binary yes/no questions at the level of the task and segment: successful completion of task/segment; within reasonable amount of time, without movement impairment; check impaired composite features. A “Yes” answer to the first two questions automatically assigned a score of 3 to the task or segment being rated. A “Yes” to the first question, with a “No” to the second provided a 2. A “Yes” to the first and second question, with a “No” to the third provided a 2 and opened up the annotation interface for composite features where at least one composite feature needed to be checked as impaired. A “No” to the 1st question provided a score of 1 and yielded a “Yes/No” question regarding completion of the initiation segment. If a “No” was provided to the initiation segment, the score became a 0. For example, in Figure 5 the clinician is first asked if the task has been performed fully. A positive answer generates the next question, which inquires if the task was performed in a reasonable time. In this instance, the clinician answers “No”, which prompts a question as to whether any movement quality elements might have impacted the task execution. Again the clinician answers “No” to this question, which generates an overall score of 2 for the task. The clinician then moves on to rating the segment movement features. Figure 3b depicts the outcome from the questions about the first segment of the task (the IPT segment). Again, the clinician answered negatively to question one and two for the segment, resulting in a generated score of 0. With this iteration, we allowed the interface to give the clinicians more freedom in assessing the rating compared to the version in Experiment 1a, but still generated the score to keep a level of consistency for the clinician. Three clinicians rated 150 tasks for Experiment 1c.

**Figure 5:** (Available per request)VAT structured decision process interface. (a) Overall video generated a score of 2 after clinician answer yes to task performed fully, and no to both whether it was performed within a reasonable time and if any of the movement qualities made the task execution challenging. (b) IPT segment generates a score of 3 after clinician selects “Task completed with no impairment”.

## Results and Discussion

Figure 6 summarizes the quantitative assessment of the three experiments. For Experiment 1a, we gave the clinicians the option to annotate the composite movement features that influenced their segment and task rating. However, the clinicians did not regularly or consistently provide these annotations. Thus IRR for composite movement features for experiment 1a is NA. There are only 3 ratings at the overall task level – the level that is most associated with the assessment of functionality. The few ratings of 3 across all experiments are associated, for more than 90% of the cases, with mildly impaired patients performing simpler tasks (tasks 1-6). Most task scores are 2s and 1s. In experiment 1b, most scores (70%) are 1s.

**Figure 6:**
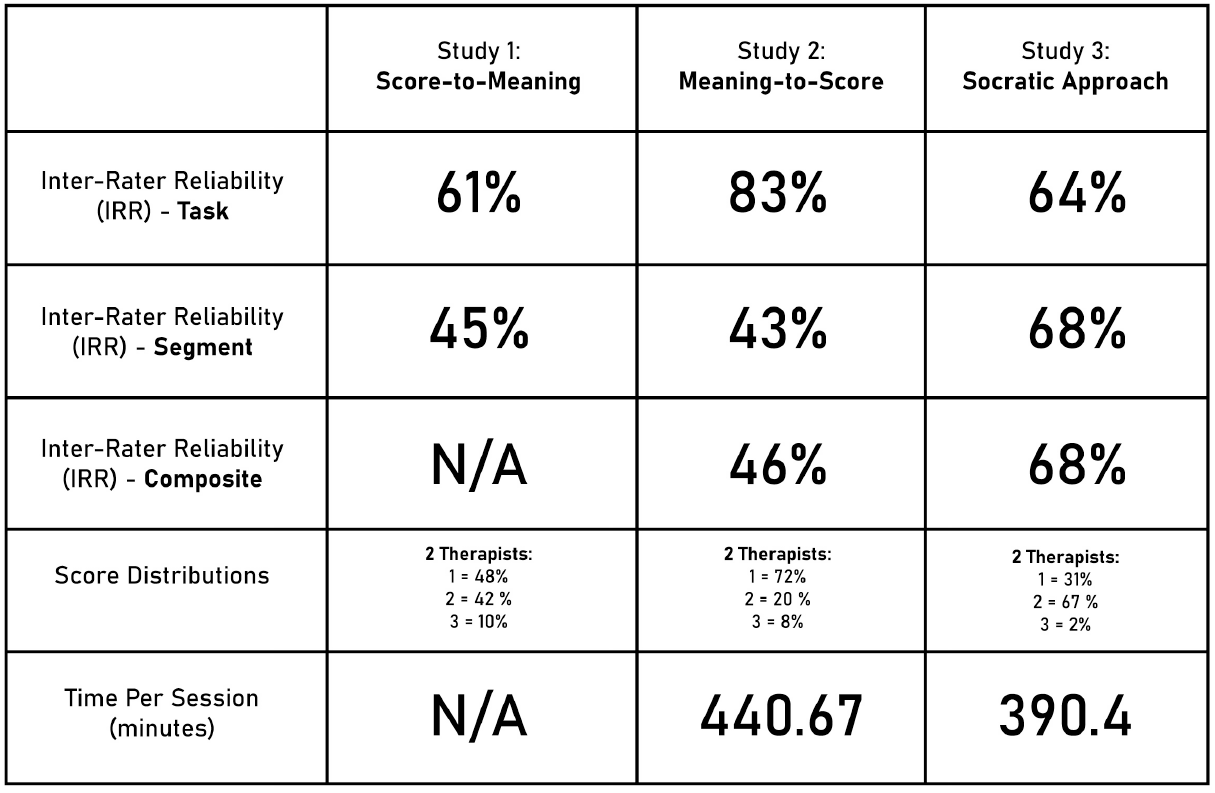
Inter-rater reliability and score distribution across all three experiments using the three version of the rating interface.

Experiment 1a produced an inter-rater reliability (IRR) of 61% on the task level, and 45% on the segment. For Experiment 1b, there was an increase in the IRR for task level to 83%. However on both the segment and composite level, the IRR remained relatively low at 43% for the segment level, and 46% for the composite. In Experiment 1c, the IRR at the level of the task dropped to 64%. However, the IRR at the level of the segment and composite levels remained high at 68%. Across all experiments, most (*>* 96%) of the rating disagreement between clinicians are *±* 1. These disagreements are fairly evenly split between ratings of 1 and 2 (one clinician gives a 1 and the other clinician(s) give a 2) and ratings of 2 and 3. As shown in Figure 6, the clinicians learned to use the interface faster when the rating process was guided/structured by the interface (Experiments 1b and 1c).

In Experiment 1a, clinicians were not asked to explicitly denote the movement quality elements that influenced their assessment of functionality, which meant that there was little agreement and standardization of assessment. The task and segment scores in Experiment 1b were directly influenced by the movement quality observations of the clinicians (if more than one movement quality element is checked as impaired then the segment score automatically becomes 1). This resulted in a radical compression of functionality results (70% of tasks are rated 1). Much of the nuance of a 4 point rating scale was lost in this case and the exploration of the relation of movement quality to functionality becomes difficult. The structured decision tree approach of Experiment 1 produced the most consistency across task, segment and composite movement feature assessment and was thus the most promising approach for exploring the relation of movement quality to functionality in a standardized manner.

Clinicians can readily embrace and utilize a top-down hierarchical approach to rating that consists of three layers: task layer (which is strongly associated with assessment of function), the segment layer (partly associated with function and party with movement quality) and the composite feature layers (associate with movement quality). This may denote that clinicians already use tacit and personalized hierarchical approaches to rating. But even when using a highly structured approach, as in Experiment 1c, clinician approaches to the impact of movement quality on functionality have similarities and differences. Figure 7 visualizes some of these differences for Experiment 1c. We show the percentage of times that a clinician checked a movement element as impaired enough to influence function when this movement feature was available for clinicians to check as impaired. For each of the times a clinician checked an available movement quality feature as impaired, we also searched to see if the other rating clinician checked the same feature for that same task. Clinicians consistently agreed on the impact of movement quality impairment on functionality for only five of the twenty available movement quality features and partially agreed on seven more.

**Figure 7:**
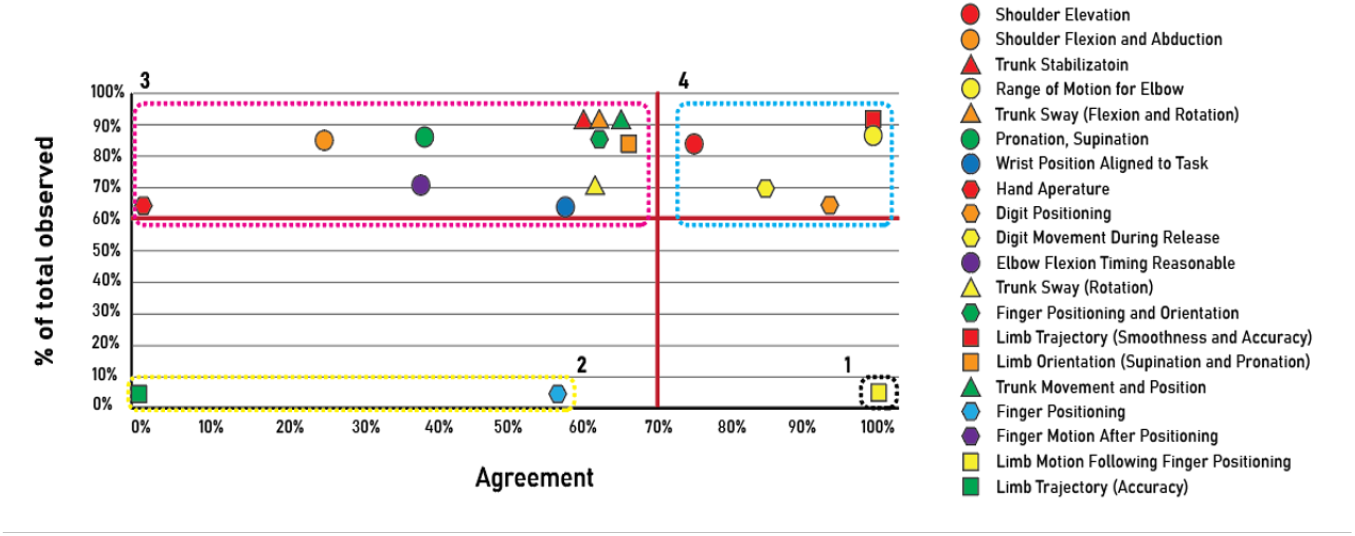
Percentage of agreement vs. percentage of observation per feature using the two clinician ratings from Experiment 1c. In this case we have set the thresholding based on the *x*-axis, meaning when both clinician check the same feature (they agreed these features are influencing functionality).

To further explore the clinician differences, in Figure 8, we visualize the level of disagreement between two clinicians as a function of the rating score. The *x*-axis spans the rating scores from severely impaired (score of 0) to unimpaired (score of 3) at the task and segment layers. As clinicians only check the composite features that relate to impaired movement for a particular segment, we use the segment ratings on the *x*-axis for the composite feature plot in Figure 8 (C). The level of disagreement is presented on the *y*-axis ranging from 0 units (agreement) to 2 units of disagreement on the task and segment level (Figure 8 (A) and (B)). For the composite layer, the disagreement is the average difference between the number of composite features checked as impaired. The pixel intensities as shown by the color bar indicates instances of different units of disagreement. Both clinicians’ ratings are used as ground truth and average instances of the level of disagreement are shown in the image. Two points emerge from the visualization: (i) most of the task and segment ratings are a score of 2, and (ii), most of the disagreement is *±*1 unit and is at its highest for rating scores of 2.

**Figure 8:**
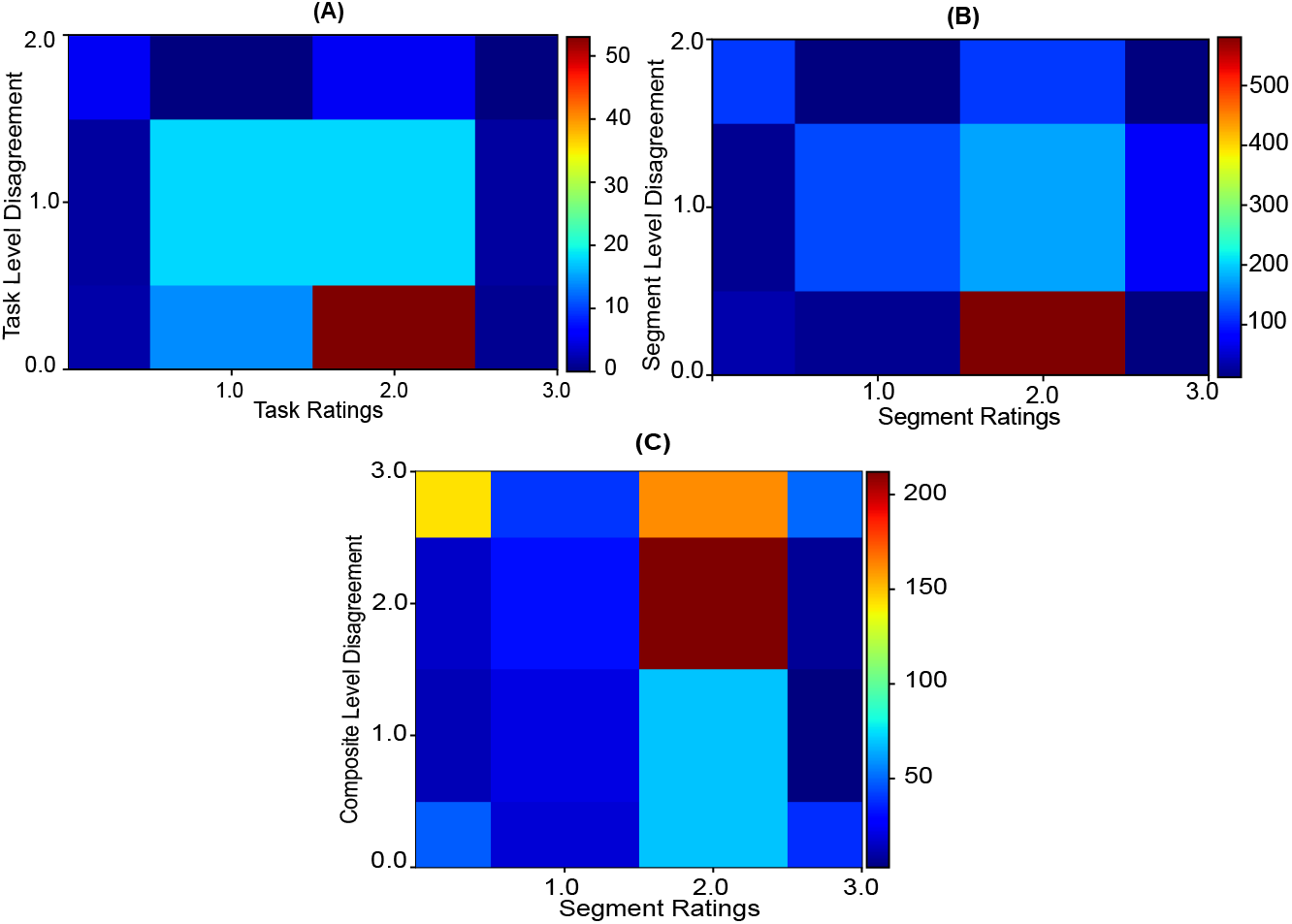
Rating scores vs. level of disagreement between two clinicians on the task, segment and composite feature level; on the x-axis ratings (0, 1, 2, and 3) are shown and on the y-axis the level of rating disagreement (0, 1, and 2 units) are shown; the color bars show the instances of any particular case.

In all experiments, clinicians are rating impaired movement since all patients have mild to moderate impairment. The clinicians can clearly separate functional movement with even slight impairment from non-impaired functional movement as shown by the high number of 1s and 2s and low number of 3s in our experiments. However, clinicians differ in their assessment of impact of movement quality on functionality. They partly differ on the movement impairments they focus on during assessment, and partly on whether the observed impairments would have a mild, moderate, or significant impact on functionality (i.e. whether an observed movement quality issue should generate a 1 or a 2 or 3 rating at the level of task performance). This variation of approaches among clinicians generates different profiles of uncertainty at each of the layers of the hierarchy. As we go down the hierarchy, the granularity increases and therefore we expect proportional division in the probability space. As a result, the span of the uncertainty decreases and the magnitude increases. As evident from Figure 8, the center of the oscillation in the task level is between scores 1 and 2 and the magnitude range is *±*1. If we go down one layer, the span of the center is narrowed around 2 and the magnitude range is *≤* 2. In the composite layer the oscillation is more narrowed around 2 but the range of oscillation is now *±*3.

We propose that relations of movement quality to functionality are probabilistic with uncertainty distributions peaking around the middle of the impaired to unimpaired continuum. By collecting more data through the interface used in Experiment 1c we can continue to inform and reveal these statistical relations. It is also important to introduce computational means that decrease the magnitude of the uncertainty as we go down the hierarchy. We propose to use an HBM to connect, in a conditional manner, the three layers of the clinicians’ ratings to a lower layer of kinematics. We propose to use the results to automate a hierarchical assessment of movement that produces interpretable results; where the functionality score is correlated with specific movement quality issues. We present our two step approach to this challenge in the next two experiments.

## Experiment 2: Computational Analysis

### Methods

The realization of the proposed interpretable automated movement assessment first of all requires a computational engine that can reliably perform the operations that clinicians readily do: automated differentiation of completed task/segments from non-completed; and automated differentiation of impaired movement from unimpaired movement. Of course this also means that the engine needs to perform automated segmentation of the captured videos. In a previous publication [22], we presented our work in developing a machine learning ensemble that could perform automatic segmentation and assess task and segment completion when analyzing the impaired patient movement used for the clinician rating experiments. In this section, we discuss how we have now extended this engine to perform these tasks across impaired and unimpaired movement so that we can begin the process of automated differentiation of impaired from unimpaired movement. For impaired movement samples, we use the patient data used for the clinician ratings, and for unimpaired samples, the unimpaired data captured in our lab (see methods section).

#### Experiments with impaired patient data

Our computational analysis solutions are developed with the goal of leveraging the hierarchical movement analysis structure and ratings provided by the clinicians to achieve a robust and interpretable automated assessment when working with limited, noisy, and variable rehabilitation movement data. In Fig. 9, we summarize the different ML engines used in our approach and how we have leveraged them to create an ensemble model for segmentation and task completion prediction. We also show the interconnection between the computational engine and the human (clinician and patient).

**Figure 9:**
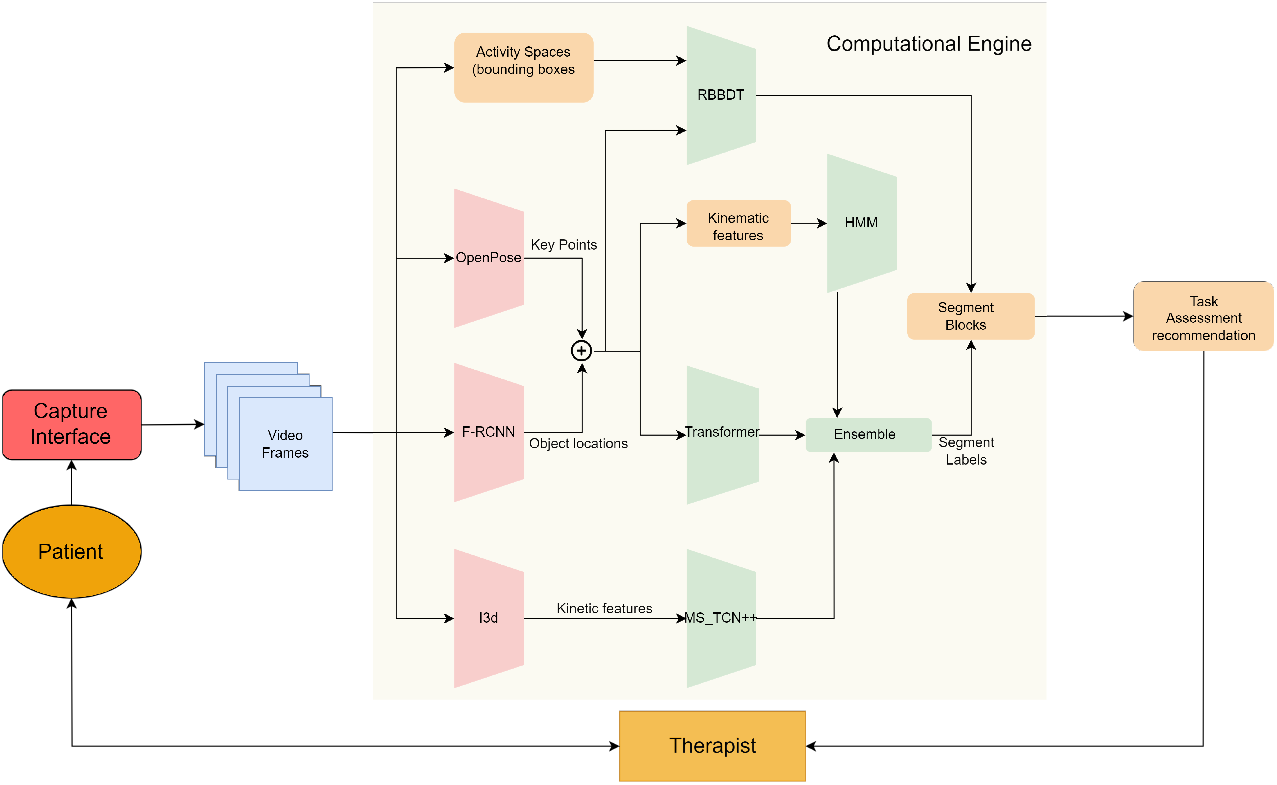
Updated block diagram from [22]. We have included the patient and the therapist blocks to complete the cyber-human loop.

Using a custom capture interface [16], we capture high quality videos and extract RGB images. As discussed in [22], the captured data was activity space variant. To reduce the variance, we also adapted a normalisation method, which is also detailed in [22]. We extract composite features from raw RGB images to train a MSTCN++[24] temporal segmentation model that uses multistage temporal convolutional neural networks. The next constituent member of our ensemble is based on the recently proposed transformer architecture which has found great success in natural language processing and time-series processing applications. We train our transformer on the keypoints extracted from the videos through OpenPose [25] and the object locations obtained using a Faster R-CNN object detection model. The last member of our ML ensemble is a Hidden Markov Model(HMM). We train our HMM using a collection of six kinematic features compiled from upper torso and upper extremity keypoints obtained via OpenPose and object locations extracted using Faster-RCNN. We implement a rule based binary decision tree (RBBDT) for segment blocks identification from per frame labels generated by the ensemble model. We leverage a unique set of features to interpret and find the transition between two segment blocks.

In experiments detailed in [22], using this ensemble model with patient data we achieved a mean performance of 85.1% with a standard deviation of 2.14% across 5 experiments using pre-defined random train-test splits of data. Using a combination of the frame level ensemble model and the segment block and task assessment algorithms using RBBDT that emerged from our work with clinicians, we are able to correctly assess segment completion about 99% of the times and task execution over 92% of the time. This means we can correctly identify which segments were completed, if those segments were completed in the right order for the satisfactory execution of a task, and whether that execution was completed within a reasonable (functional) amount of time.

#### Experiments with unimpaired subject data

The patient data used for these earlier experiments has very few unimpaired movement examples (very few examples rated a 3). For our machine learning ensemble to be generalizable and scalable, it needs to predict the edges with precision. We can add edges to the existing probability space by introducing training tasks performed by unimpaired subjects. We removed all incorrectly performed tasks from the the data we collected with unimpaired subjects so as to be able to assign a rating score of 3 to all tasks performed by unimpaired subjects that we included in these experiments. The ability of our ensemble to segment training tasks and assess task and segment completion across impaired and unimpaired data is critical for training the ensemble to automatically differentiate impaired from unimpaired movement and, in the future, automatically extract correlations of movement impairment to functionality.

We performed three experiments using mixes of impaired patient data and unimpaired subject data and compared the results to the experiments from [22]. The data mixes of the three new experiments are shown in 1.

### Results and discussion

In Table 2, we show the comparative analysis for frame wise label prediction for Experiments 2a, 2b, and 2c from Table 1. As evident from the results, Experiment c has the highest accuracy for per frame label prediction. This supports our argument that training the models with both impaired and unimpaired data makes the model more generalised and also increases the performance of the model compared to [22]. In Experiment 2a, the models were trained using noisy patient data captured in a variant activity space. However, we evaluated using unimpaired subject data captured in an invariant space. As shown in Table 3 the models achieved 88.5% accuracy in segment detection of unimpaired subject data even when the model is trained using only noisy patient data. It is evident that the model is learning the relationship between movement patterns and the function(s) embedded in each training task. In Experiments 2b and 2c, the models were trained once using 75% patient data and 25% unimpaired subject data. However, for Experiment 2b, the test set was the whole patient dataset and for Experiment 2c the test set included both patient (60%) and unimpaired subject (40%) data. The target of these two experiments was to improve the model performance by increasing the size of the dataset and by introducing a different domain data (unimpaired subject data). In both of these cases the overall performance improved significantly as shown in Table 2 and 3 .

**Table 1:** Experiments with Unimpaired invariant data

|  | Train | Test |
| --- | --- | --- |
| Experiment 2a | trained model using impaired | unimpaired data |
| Experiment 2b | 75% impaired 25% unimpaired | test set using impaired |
| Experiment 2c | 75% impaired 25% unimpaired | 60% impaired 40% unimpaired |

**Table 2:**
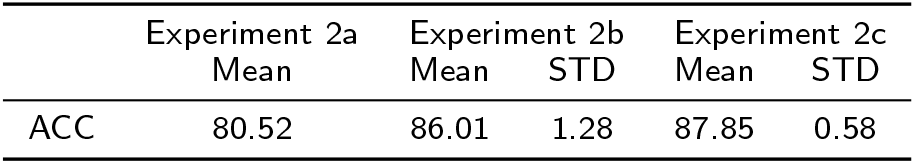
Frame Wise Segmentation Results for Experiments 2a, 2b and 2c in Table 1

**Table 3:** Segmentation Block Results for Experiments 2a, 2b and 2c in Table 1

|  | Experiment 2a | Experiment 2b |  | Experiment 2c |  |
| --- | --- | --- | --- | --- | --- |
|  | mean | mean | std | mean | std |
| Segment accuracy | 88.52 | 98.71 | 1.1 | 98.46 | 1.6 |
| Precision | 84.13 | 98.65 | 1.1 | 98.22 | 1.7 |
| Recall | 83.38 | 97.92 | 1.7 | 97.63 | 2.01 |

Our expanded ensemble model is able to reproduce the unimpaired movement scores (3s) on the tasks and segment level with over 90% accuracy. In Figure 10, we show the comparison between the computer generated task scores and the clinicians’ ratings for 440 videos that combine videos of patient performed tasks from Experiment 1b and 1c and videos of task performance by unimpaired subjects. The ensemble can correctly label with a score of “3”, 130 of 140 videos that had no impairment or had minimal impairment and had thus received a rating of 3 by the clinicians. In the rating rubric, a score of 0 means the patient didn’t attempt the task or complete the first segment(IPT). A score of 0 doesn’t reveal the relation between movement quality and functionality and therefore, we use the RBBDT component of the ensemble to reliably and automatically reproduce all the 0s.

**Figure 10:**
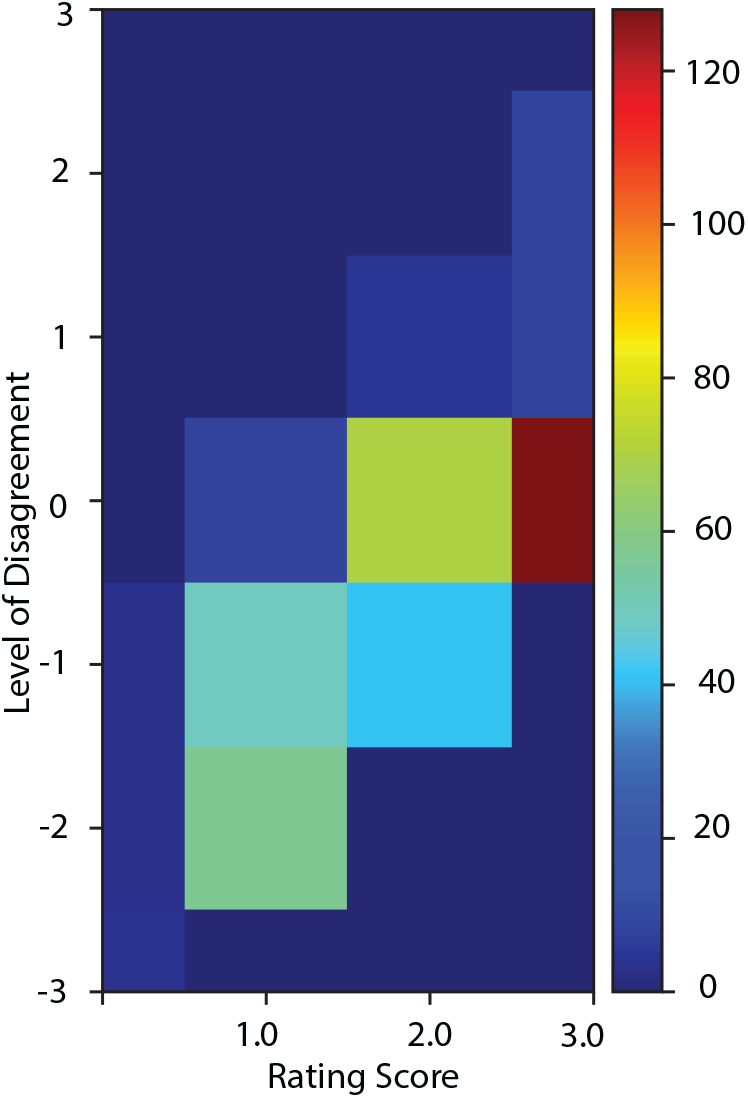
Level of disagreement vs rating scores between the computer generated task scores and the clinician’s ratings. On the y-axis the level of disagreement is shown. The 2-D pixels indicates the instances of any particular case and the value of the pixels are indicated by the color bar on the right.

**Figure 11:**
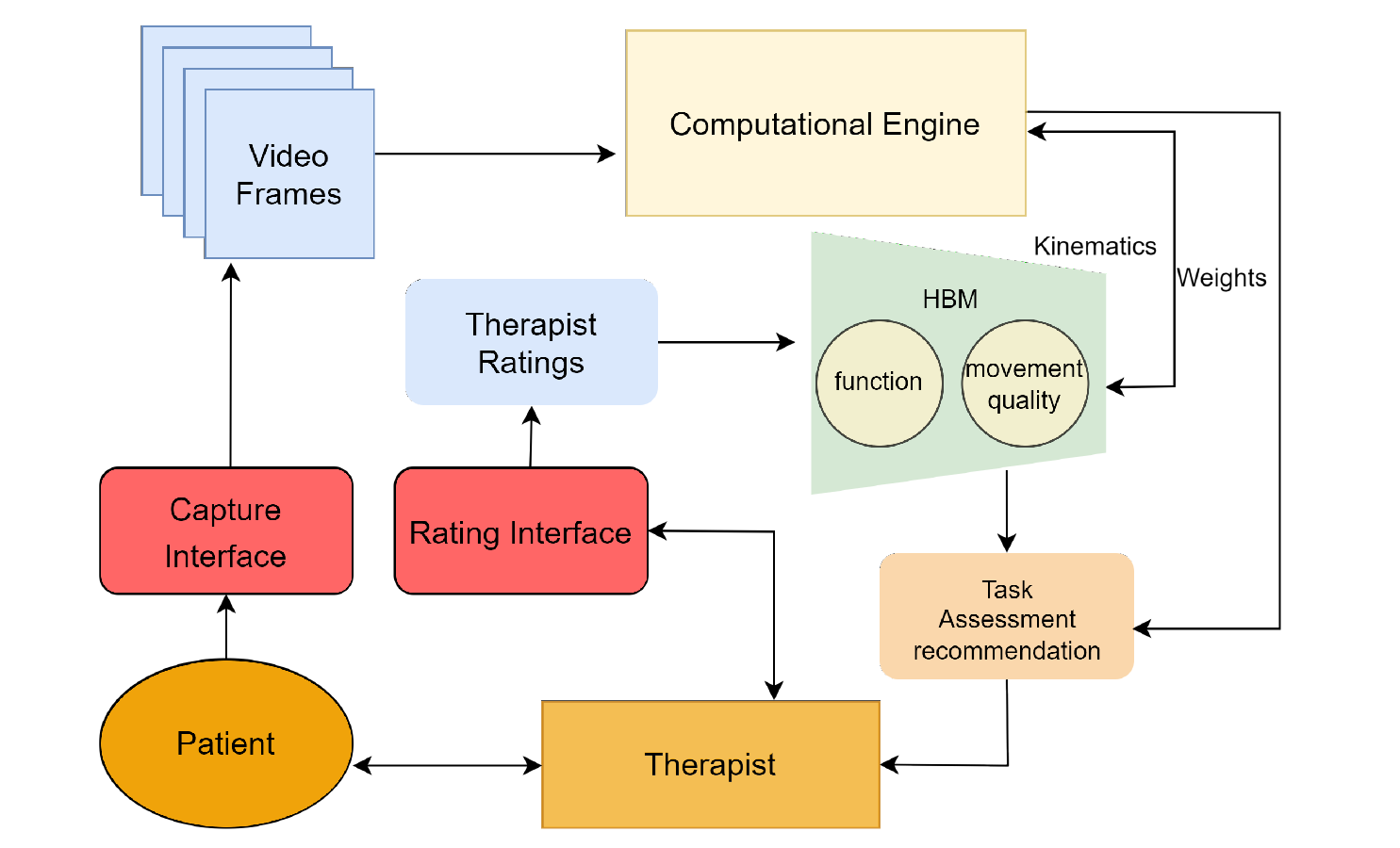
Block diagram incorporating the HBM with the computational engine

However, the computational ensemble cannot reliably produce automatically the 1 and 2 ratings provided by clinicians. Separating a rating of 2 from a 3 or a 1 requires a detailed understanding of movement quality, and their relation to function. As discussed in Experiment 1c, there is significant uncertainty even among clinicians when using ratings 1 and 2. Clinicians reduce this uncertainty by using tacit and explicit knowledge to calculate the probable effect of an observed movement quality issue on task dependent functionality. In the next section, we show how an HBM can capture and reproduce the probabilistic process used by the clinicians and connect this process to computationally extracted kinematics. The HBM can then be integrated into the computational ensemble, thus, allowing the ensemble to automatically and reliably produce all four possible ratings (0-3), at the task and segment level, and denote potential movement quality issues that may be significantly impeding functionality and require focused therapy.

## Experiment 3: Formation of the HBM

### Methods

We aim to develop a model that can integrate raw features and kinematics extracted computationally in the hierarchical and statistical assessment structures of rehabilitation movement used by clinicians. Our goals are to help quantify relations of movement quality to functionality and advance automated interpretable analysis of rehabilitation movement. Hierarchical models are used in many other areas of structured human activity such as language [26], music [27], logic and computation [28], sports [29] and motor learning in general [30]. This has inspired the development of generalizable models of human intelligence [31], [32] and human learning [33]. Tenebaum et al [21] suggest that human performance in complex situations with noisy data can be modeled as a hierarchical Bayesian model (HBM) [21]. Building on this prior work, we propose an HBM for integrating the information captured by the expert ratings of videos with a hierarchical computational analysis of movement using combinations of knowledge-driven and data-driven algorithms. As we show below, using the HBM we can begin to quantify the relations of movement quality to functionality even when using limited and noisy data. We can integrate the HBM in our computational ensemble and start real world automated assessment applications in the clinic and home with limited training data. As additional data becomes available through these applications, the results will become more robust.

#### Expressing HBM layers using clinician ratings

The hierarchy layers of the proposed HBM have already been defined by our prior work with expert clinicians: task performance, segment performance, composite movement features, kinematics, and raw features as depicted in Figure 1. In Experiment 3 of this paper, we want to begin to quantify the statistical relationships of these layers. We first exploit the expert knowledge captured through the rating rubric and clinician ratings to express the relationships of layers as conditional probabilities. The conditions are imposed on the prior level of the hierarchy to get a posterior of the immediate lower level. The mathematical notations used to calculate the probabilities between each layer are given in Table 4.

**Table 4:**
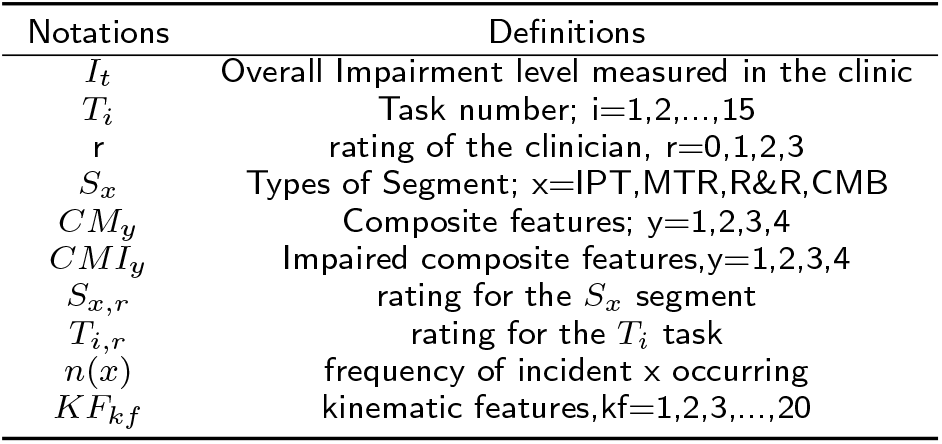
the mathematical notations and their definitions used in the formulation of HBM

By expressing the relationships between all the layers, we can then define a joint posterior probability to reveal which changes in movement quality (*CMI_y_* to *CM_y_*) have the highest probability of affecting function (raising *S_x,r_* and/or *T_i,r_* to 3) as well as the *CMI_y_, S_x_, T_i_, r* sets where this is more observable. We can then explore the relationship between the most impactful *CMI_y_* to *CM_y_* changes in movement quality to the kinematics data being captured through low cost/low intrusion infrastructure (i.e. video camera and/or IMUs). We use the clinicians’ ratings to compute the conditional probabilities up to the composite movement features layer of the hierarchy. We then replace the composite features layer with the raw kinematics layer, thus deriving conditional probabilities relating task and segment performance to kinematics. We then use a novel matrix structure to combine the conditional probabilities and compute the relationships between impactful changes in composite features and correlated changes in kinematic features. This makes possible the automated detection of movement quality issues that may be effecting task performance and overall functionality and sets the stage for intepretable automated assessment of rehabilitation movement.

#### Task-segment relations

To investigate meaningful relations of task (x) and segment (i) execution we need to calculate conditional probabilities for all the (*x, r*)*and*(*i, r*) pairs for any given clinician’s rating (r). The overall space of possible relations between the Task layer and Segment layer can be divided into three categories of task-segment completion relations

- complete execution in both layers, *P* (*S_x,r_ ≥* 2*|T_i,r_ ≥* 2)
- incomplete execution in both layers, *P* (*S_x,r_ ≤* 1*|T_i,r_ ≤* 1)
- incomplete execution in one of the two layers, *P* (*S_x,r_ ≥* 2*|T_i,r_ ≤* 1) or *P* (*S_x,r_ ≤* 1*|T_i,r_ ≥* 2)

Complete tasks and segments may be rated 3 or 2 based on movement quality observations by the clinician. Therefore each of these three categories of task-segment completion relations presents three subcategories of relations: two 3s, two 2s, a combination of one 2 and one 3. We will now briefly discuss each of the categories (and their subcategories) and calculate the conditional probabilities for each. All the frequencies are calculated using clinician ratings of the segment and tasks.

##### a) Complete execution in both layers, P (S_x,r_ ≥ 2|T_i,r_ ≥ 2)

When all the segments are executed (with or without impairment) and the task is completed within the allowable time, the movement quality observations of the clinician can create the following rating combinations:

***i) Unimpaired execution in both layers,*** *P* (*S_x,r_*_=3_*|T_i,r_*_=3_). The probability of unimpaired execution at the task level (r=3) being related to unimpaired (r=3) execution at the segment level for all pairs of relevant *S_x_, T_i_* would be calculated using,

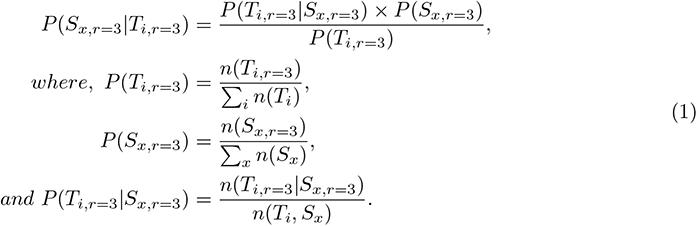

***ii) Impaired execution in one of the layers,*** *P* (*S_x,r_*_=2_*|T_i,r_*_=3_) ***or*** *P* (*S_x,r_*_=3_*|T_i,r_*_=2_). The type of segments that have a higher probability of receiving a 2 when the task receives a 3 (the (*x, r* = 2), (*i, r* = 3) pairs) would denote segment execution where the movement impairment is not significantly influencing function.

***iii) Impaired execution in both layers,*** *P* (*S_x,r_*_=2_*|T_i,r_*_=2_). The type of segments that have a higher probability of receiving a 2 when the task receives a 2 (the (*x, r* = 2), (*i, r* = 2) pairs) would denote the segments we would need to focus on in terms of movement changes that affect function. Therefore, we would need to calculate

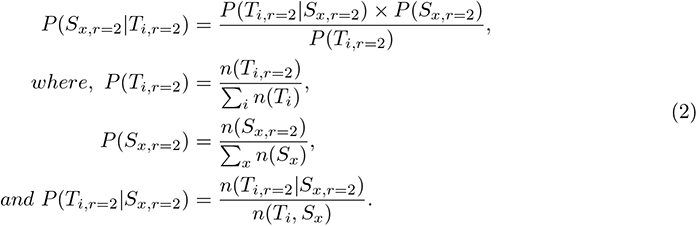

##### B) Incomplete execution in either of the layers, P (S_x,r_ ≥ 2|T_i,r_ ≤ 1) or P (S_x,r_ ≤ 1|T_i,r_ ≥ 2)

A high probability of a task receiving a *r ≥* 2 when a segment receives a *r ≤* 1 rating would denote significant compensation during that type of segment in a way that negates the full use of the affected limb. For example, a bi-manual manipulation (screwing of a jar lid) task is fully executed using primarily rotation of the unimpaired limb during the MTR stage. A high probability of a segment receiving a *r ≥* 2 rating when a task receives a *r ≤* 1 rating would denote that this type of segment is not the one causing the challenge with this task and would direct us to find the challenge in other segments.

#### Incomplete execution in both layers, P (S_x,r_ ≤ 1|T_i,r_ ≤ 1)

A high probability of a task receiving a *r ≤* 1 when a segment receives a *r ≤* 1 rating would denote significant movement impairment across that type of segment and other segments resulting in an incomplete task execution. Therefore, the relation of each individual segment to a task is not fully observable and thus not meaningful.

#### Segment-composite feature relation

The relationship between the segment layer (*S_x_*) and the composite movement feature layer (*CM_y_*) will indicate which composite features contribute to the segment’s ratings. This probability space is divided into three categories of completion-impairment relations.

Completed segment execution with movement quality impairment, P (CMI_y_|S_x,r=2_) We would calculate this by:

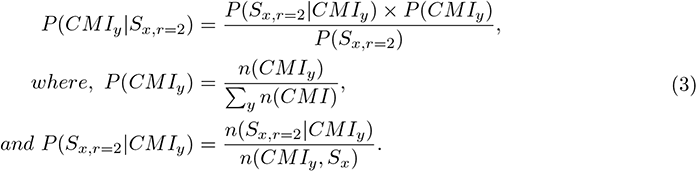

Here, the frequency of any particular impaired composite features, *CMI_y_* indicate the total number of times that particular feature, *y* was denoted by the clinician as impaired when assessing the execution of a type of segment. To show this as a ratio of the total times a feature is available for denoting as impaired we can rewrite the equation as:

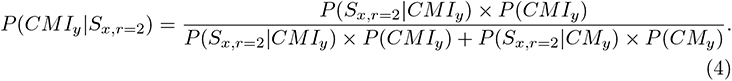

***Incomplete impaired execution,*** *P* (*CMI_y_|S_x,r_*_=1_). We can similarly calculate the probability of incomplete execution of a segment being related to the impairment of a specific composite feature using the frequencies of each incident.

***Complete unimpaired execution,*** *P* (*CM_y_|S_x,r_*_=3_) We can also calculate the contributions of composite features to the complete and unimpaired execution of a segment using:

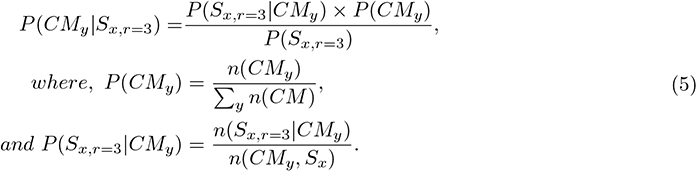

Similar to (3), the frequency of any particular unimpaired composite features, *CM_y_* indicate the total number of times that a particular feature,*y* was available for observation but was not denoted as impaired by the clinician.

#### Increasing granularity of HBM movement quality layers through computational experimentation

We can now calculate the joint posterior probabilities by taking a product of the conditional probabilities for each of the impaired and unimpaired composite features:

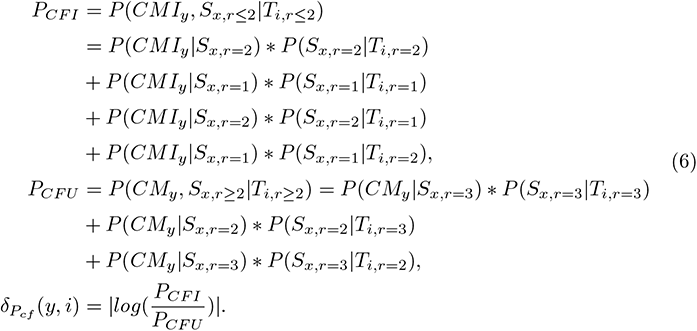

Here, *P_CFI_* and *P_CFU_* denote the joint posterior for impaired (CFI) and unimpaired (CFU) execution. We calculate all the task and segment layer probabilities for each case of CFI and then sum them together to calculate the joint posterior. We repeat the same process for CFU. We hypothesize that the more observable features have prominent relationships with all the layers and will contribute to higher probabilities. The impaired execution involves multiple cases and thus, the dimension of the feature space will be large. Therefore, we can use thresholding to rule out some of the cases with very small conditional probabilities. We then need to calculate the *P_CFI_* and *P_CFU_* delta for each composite feature, and *CF_y_* to estimate the impact of each feature in differentiating an unimpaired from an impaired execution. The *δ_Pcf_* has the dimension of (*y, i*) meaning for each task, *i* the composite features, *y* with large probabilities may have a significant impact on functionality and related task execution assessment. As we collect more data, the quantification of the impact of movement features on different types of functionality (different types of tasks and segments) will become more robust and help disambiguate the differences between clinician ratings at this layer of the assessment hierarchy. Adding a kinematics layer below the composite features layer, and connecting these kinematics to the composite features, allows us to increase the granularity of observation of movement quality and further disambiguate the relation of movement quality to functionality. Our previous work [2] and [22] established a list of twenty kinematic features that can be used to capture and analyze movement quality issues affecting functionality. However, there are currently no direct mappings of these kinematic features to composite movement features used by the clinicians for assessing movement quality and the performance of standardized segments. To establish these important connections, we first repeat the delta calculation we described for composite features for each of the computationally extracted kinematics. We consider all the probability sub-spaces where segment ratings are different and for each rating we compute the conditional feature mean, *F* using:

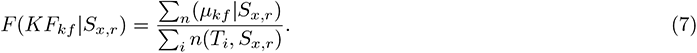

Here, *µ_kf_* is the sample mean of the kinematic feature, *kf* per segment, *S_x,r_*. We calculate the mean per data and then add all the *µ*’s for *n* number of occurrences of the case *S_x,r_*. The *F* notation is used to separate conditional probability, *P* from the conditional feature mean calculated using (7). This way we calculate all the conditional segment-kinematic feature probabilities for all the cases. Similar to (6), we can calculate the joint posterior probability and compute the *δ*.

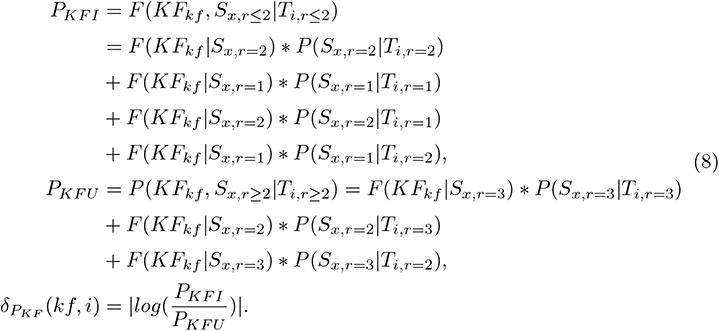

Here, *P_KFI_* and *P_KFU_* denote the joint posterior for impaired and unimpaired execution respectively. Here, the *δ_PKF_* signifies the kinematic components that change significantly between impaired and unimpaired performance of segments. This technique allows us to map performance of segments to change patterns of kinematic components and indicate what changes in kinematic components may significantly affect the execution of a type of segment. The *δ_PKF_* has the dimension of (*kf, i*) meaning for each task, *i* the kinematic components, *kf* with large probabilities are significant for integrated assessment of movement quality and functionality.

Since both of the composite movement features and kinematic component deltas have the task and segment layers as a common conditional reference, we can simply multiply them to get a (*kf, y*) matrix to define the relationship between composite features and kinematic components,

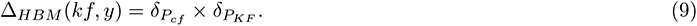

### HBM Analysis Results and Discussion

Figure 12 shows the relation of the (Δ*_HBM_*) of the 16 composite movement features to the 20 kinematics components. The color bar represents the normalised value of Δ*_HBM_* . When a clinician denotes a movement feature as impaired, we can use this table to further estimate the level of impairment by tracking differentiation from the unimpaired mean for the kinematic components strongly correlated to that particular composite movement feature. We can also estimate the effect of the denoted impairment on segment execution using the deltas of kinematic components established earlier. However, as shown in earlier sections, these relations are conditional (i.e changes in movement quality impact execution of different segments and tasks differently). Connecting the kinematics layer to the performance of composite features and segments directly (as discussed above) reveals the global movement quality to functionality relations. To also reveal the conditional relations, we need to calculate the cascading probabilities that connect task and segment execution to composite features quality and in turn kinematic patterns. The addition of the kinematics layer to the HBM allows us to increase the granularity of observation of movement quality. The conditional probabilities across the four layers of the HBM (task, segment, composite, kinematics) allow us to further disambiguate the relation of functionality to movement quality.

**Figure 12:**
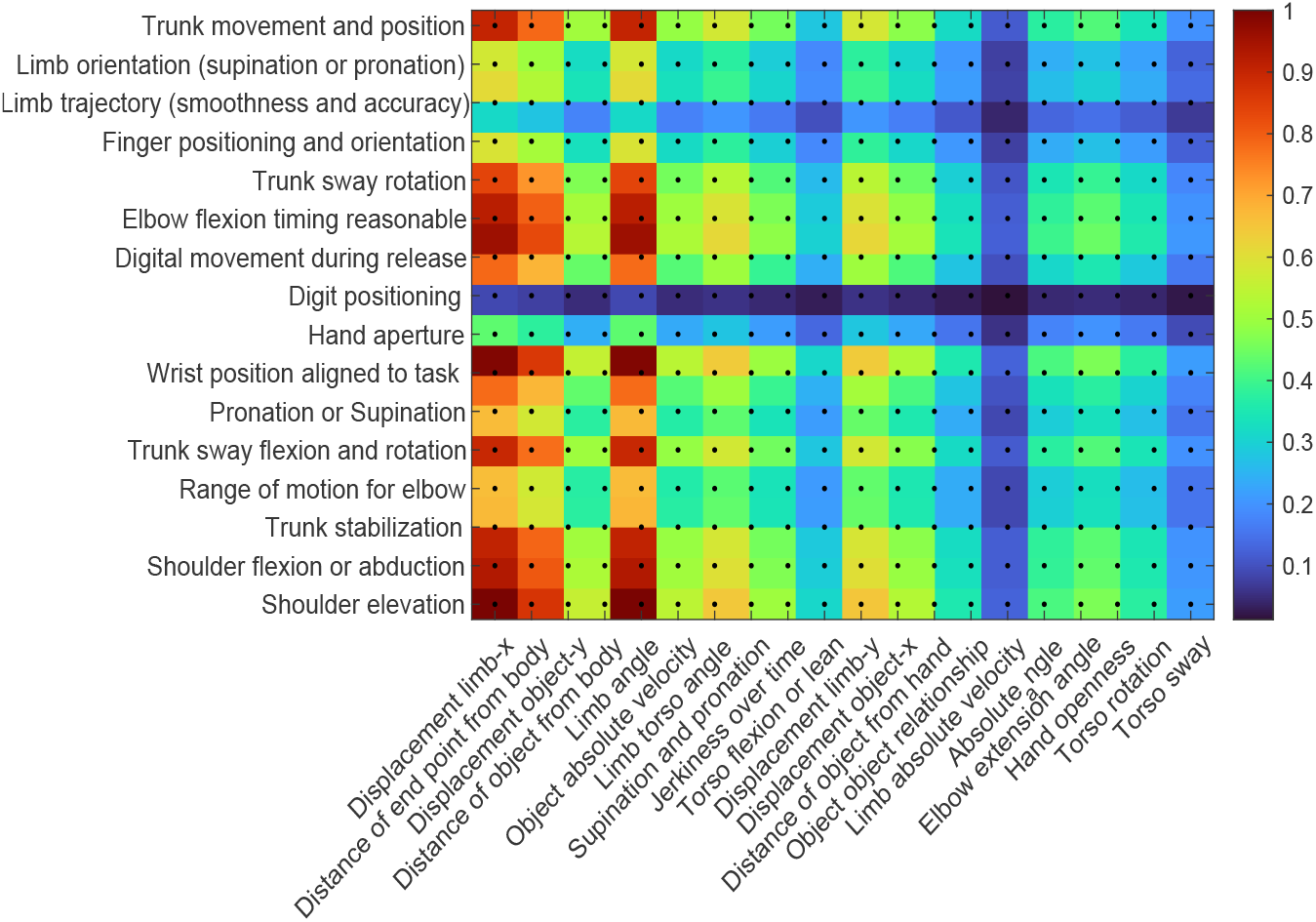
Visualization of Δ*_HBM_* feature values captured between composite features (x-axis) and kinematic features (y-axis).

For example, the four layer HBM can help us analyze the potential contributing factors for any task being rated 2. We first use the task ratings to estimate which of the segments of that task have the highest probability of impaired execution (Figure 13). We then estimate which composite features have the highest probability of influencing the performance of these impaired segments (Figure 14 (A)). We finally estimate which kinematic components have the highest probability of defining the details of each composite movement feature impairment for each of the impaired segments (Figure 14 (B)). The x-axis in Figure 14 (B), has 3 segment labels since the total time of the segment *IPT* is used as the temporal segmentation (the segment concludes at end of *T*). Also, since the computational analysis included only Tasks 1 - 10, we do not include *CMB* or *M* &*TRB* segments as these are only found in Tasks 11 and 12. For the same reason the last four composite features in Figure 14 (A) have zero instances.

**Figure 13:**
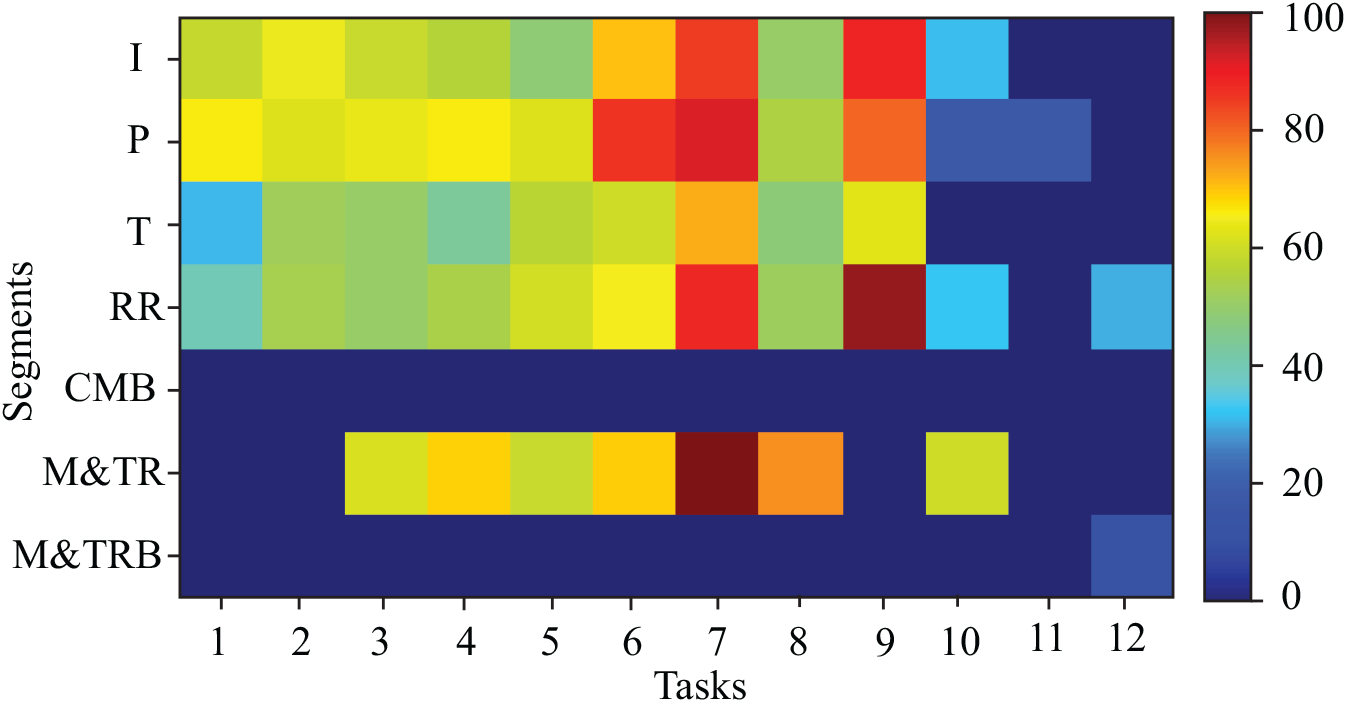
Demonstrating the probability of a segment to have a rating of 2 (impaired) when a task was rated 2.

**Figure 14:**
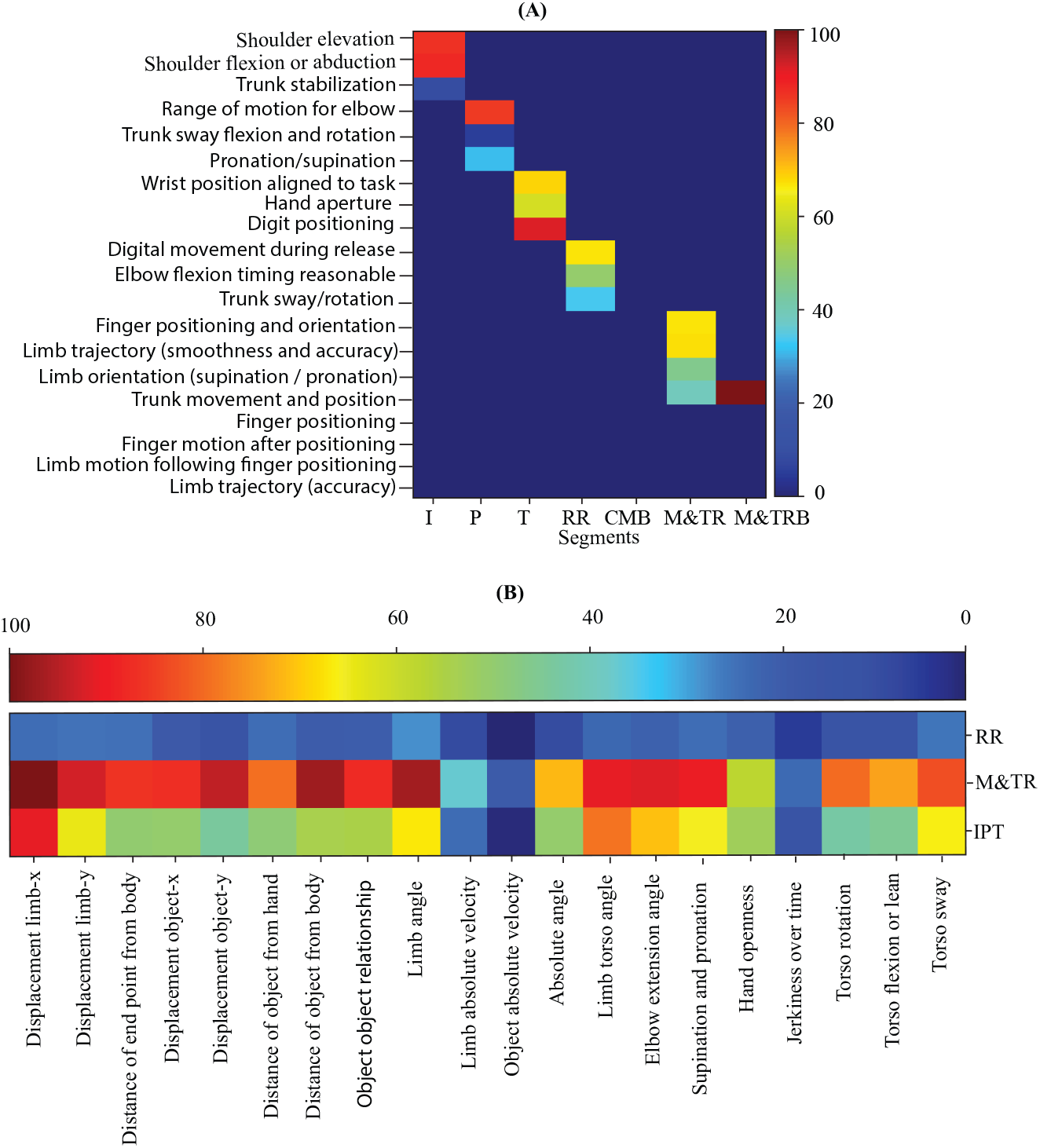
Demonstrating the probability of a composite feature (A) and kinematic components (B) to influence the performance of an impaired segment (rated 2).

Comparing Figure 14 (A) to Figure 7 shows that the inter-layer probability graphs produced through the HBM fully align with clinician observations. The five movement quality features that are most often observed by the clinicians when rating videos are clustered in the upper right hand corner of the Figure 7. This cluster has significant overlap with the cluster of features showing the highest probability of affecting function (performance of segments) in Figure 14 (A). Therefore, the four layer HBM can be used in a top-down manner (task to segment to movement features) to reveal and quantify (in a statistical manner) the relations of functionality and movement quality that emerge from clinician ratings. The HBM can also be use in a bottom-up manner (from the kinematics upwards) for the automated calculation of the relation of movement quality impairment to functionality. Figure 15 shows that we can calculate the probability of a particular composite movement feature influencing the performance of a task. Figure 16 shows that we can calculate the probability of a particular kinematic feature influencing the performance of a task.

**Figure 15:**
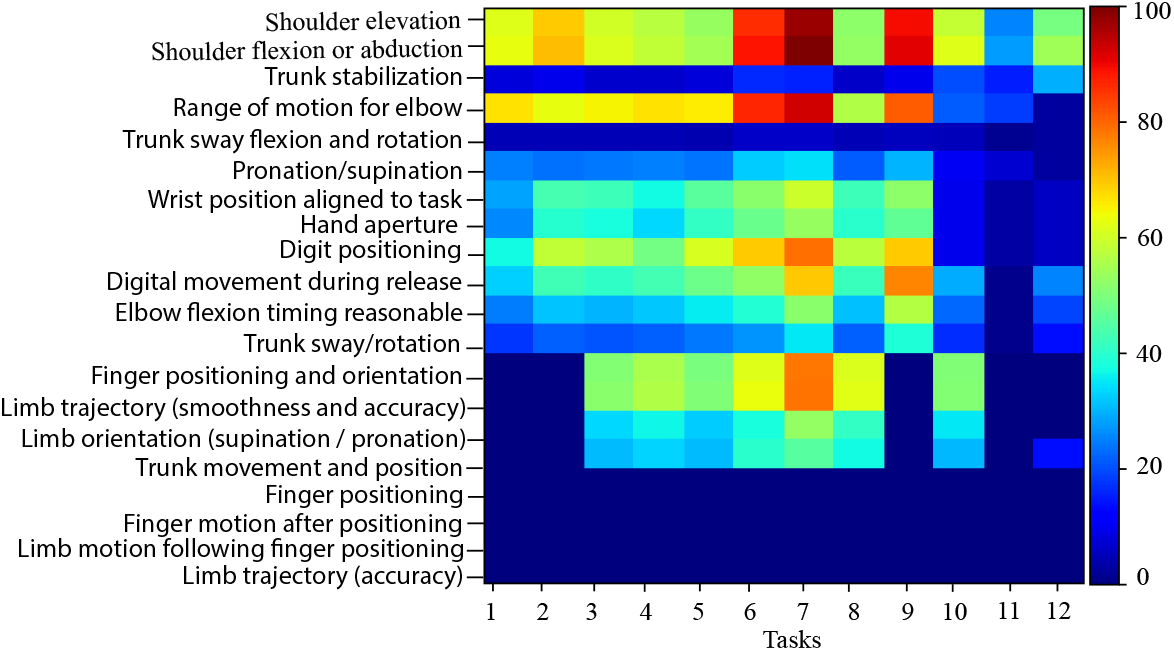
Demonstrating the probability of a composite feature to influence the performance of an impaired task (rated 2).

**Figure 16:**
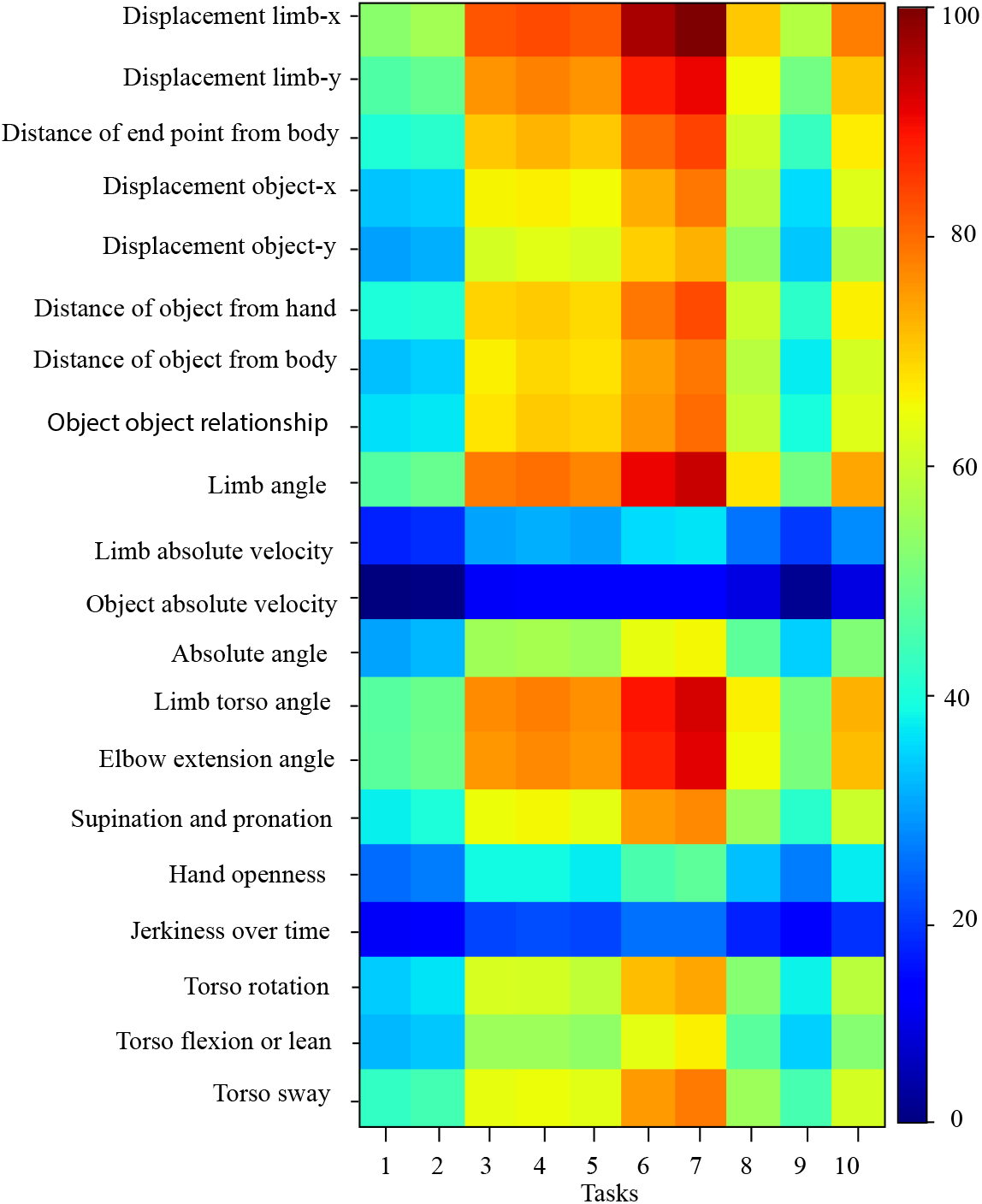
Demonstrating the probability of a kinematic feature to influence the performance of an impaired task (rated 2).

**Figure 17:**
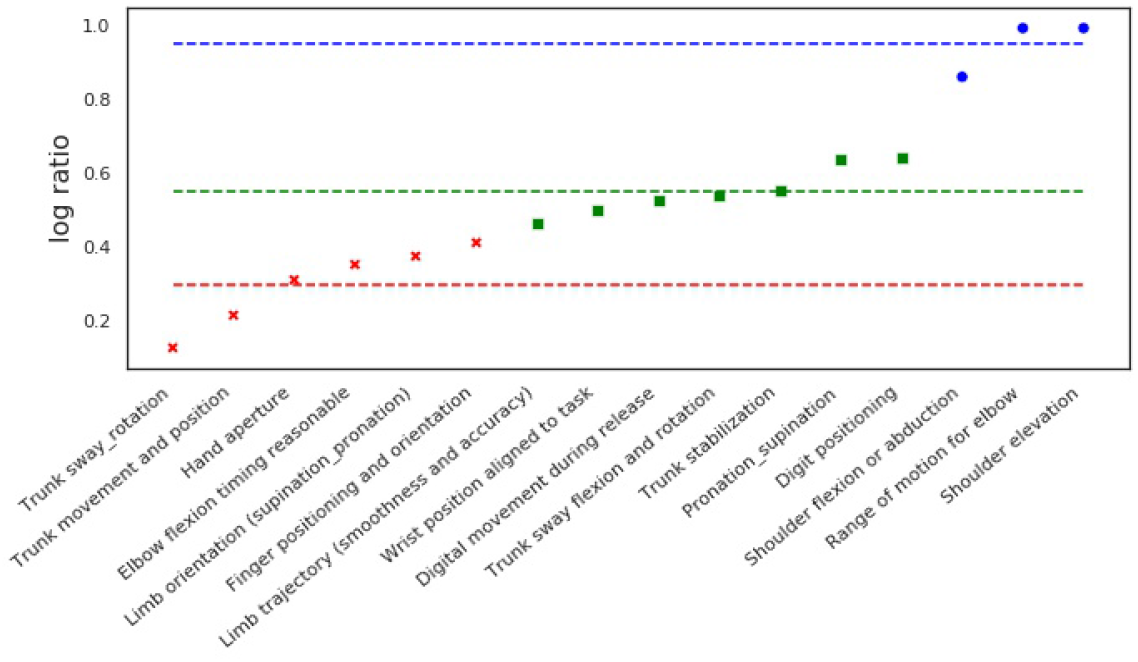
Visual representation of the three different clusters formed for limb angle.

### Automatic Assessment: Weight Optimization and Transformer

One of the goals of the proposed approach is to integrate the HBM into our computational ensemble in order to be able to perform semi-automated and interpretable assessment of rehabilitation movement. To achieve this goal we need to appropriately weight the raw kinematic features before giving them as inputs to the machine learning models. The transformer model or the HMM models will then be optimized to predict the correct segment labels and ratings with the weighted features as input. While there could be multiple ways of producing weights, we adopt an interpretable approach for the initial weight generation. Specifically, we use an unsupervised clustering algorithm to cluster the raw features into three categories: (i) mild, (ii) medium and, (iii) severe cases of movement impairment. This step is carried out for every raw feature independently.

Concretely, consider the final feature matrix Δ*_HBM_* given in (9) that captures the relationship between (i) kinematic features and (ii) composite features. Let Δ*_HBM_ ∈* R*^N×M^* . We run k-means clustering for all columns and rows independently i.e., say for each kinematic feature, we take the features corresponding to all *M* composite features and cluster them into three clusters as described. The cluster centroids are then used to weight the features. In Figures 20 and 18, we plot the clusters and cluster centroids for 4 randomly chosen kinematic and composite features. We observe that in most cases, the features can be clustered into three distinguishable categories thus making it amenable to produce meaningful weights. We can then use the centroid of these clusters as the weights of the input data to our computational ensemble (the Transformer, MSTCN++ HMM) so as to:

i. compute quality of performance of composite features from kinematic features/ raw visual features
ii. rate segments automatically
iii. rate tasks automatically.

**Figure 18:**
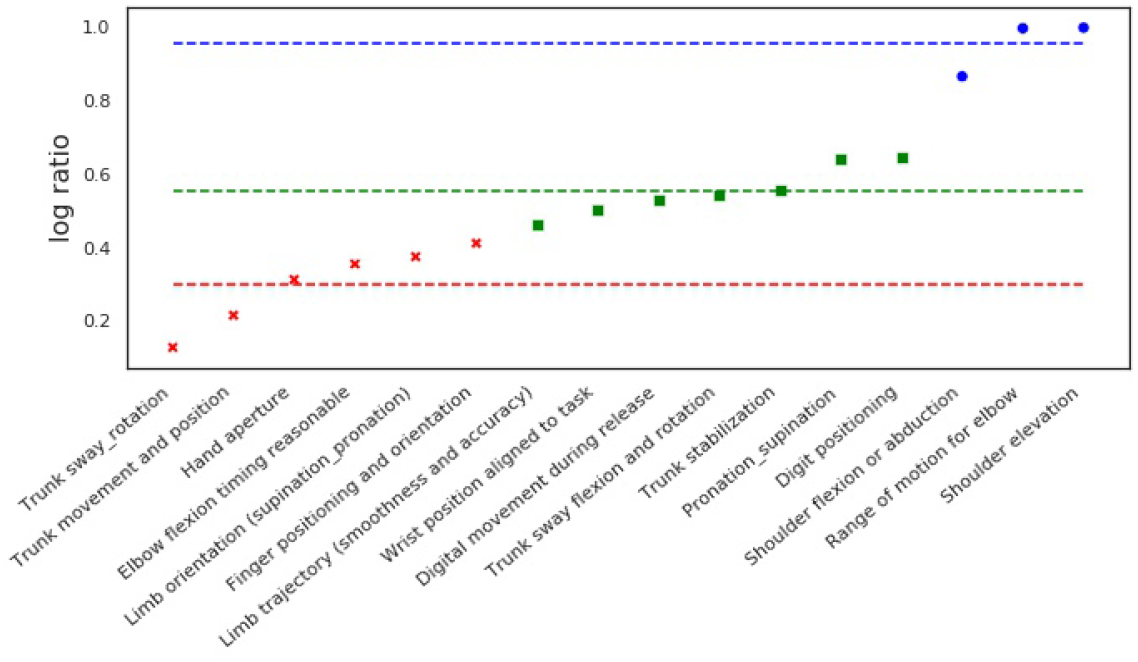
Three different clusters for the feature: displacement of limb in the x direction.

**Figure 19:**
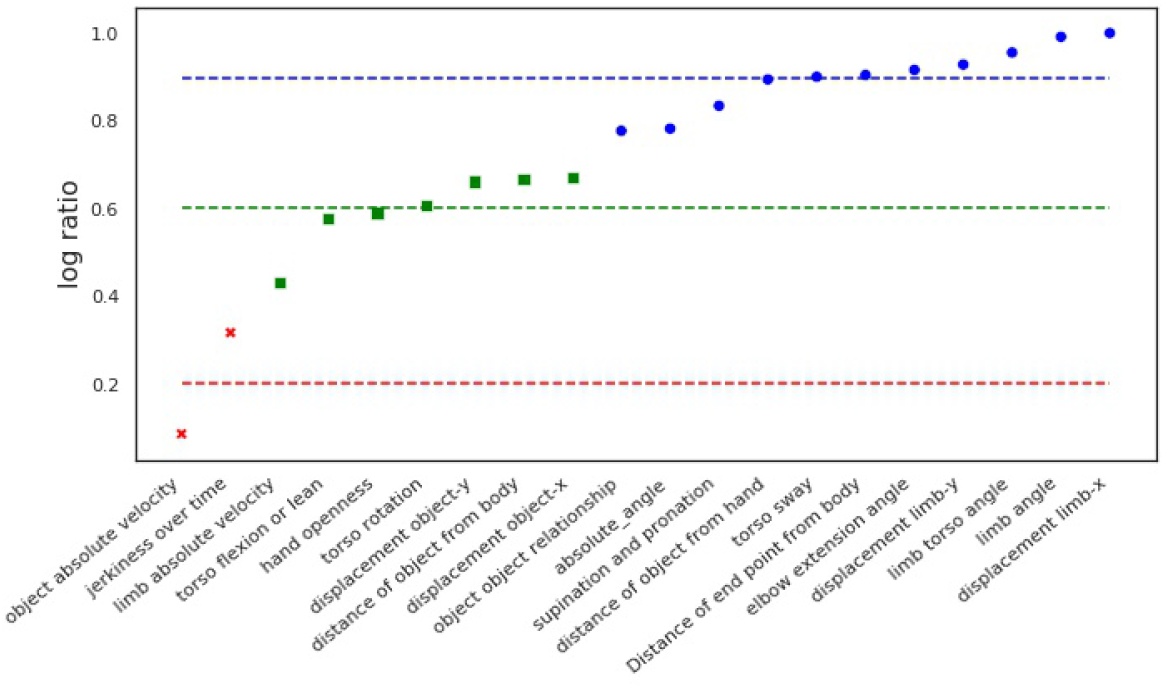
Three different clusters formed for the feature: shoulder elevation.

**Figure 20:**
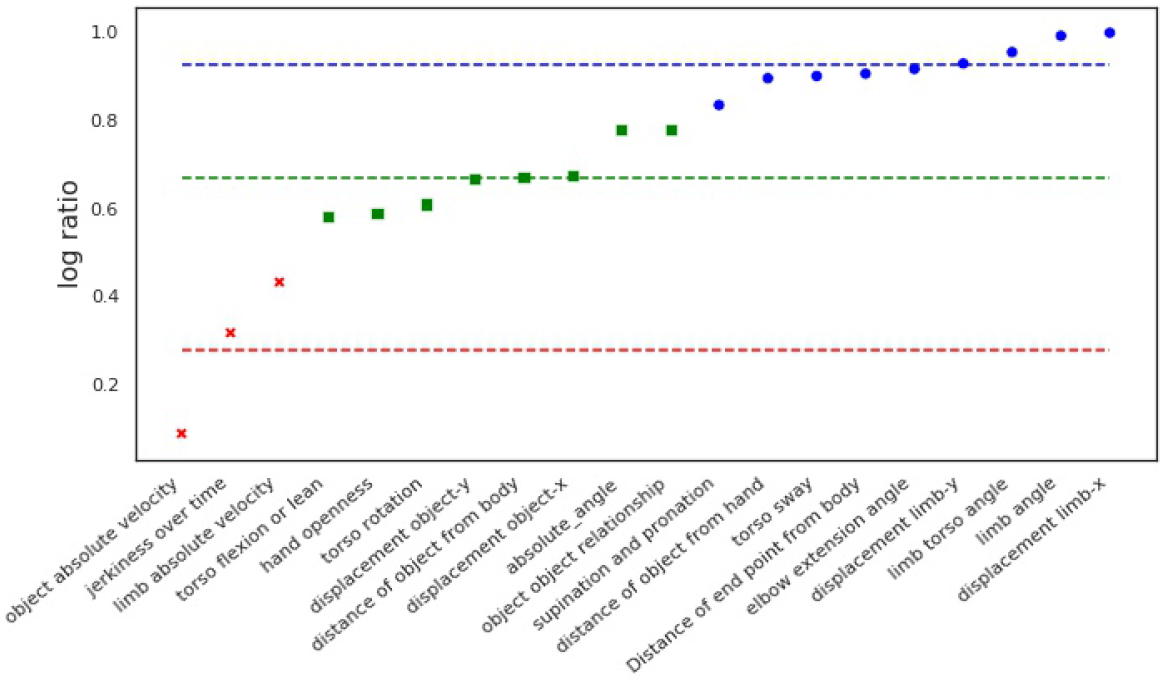
Three different clusters formed for the feature: range of motion for elbow.

We can then adjust our ensemble model to produce a joint rating of segment blocks across the HMM, Transformer, MSTCN++ and other algorithms. We can then rerun the HBM pipeline for the computational results to explore if there are significant differences. Some of these potential differences may be due to elements that are observable by data driven algorithms but not captured in the rating rubrics created by the expert clinicians. For example, there could be a composite movement feature that is important for a particular segment/task pair that was not included in the rating rubric. We can bring this information back to the expert clinicians to continuously improve the rubric and rating interface. As we capture more varied data and the HBM is more generalized, then we can start tuning the weights. The HBM weights could also be used as input to other machine learning algorithms (beyond the HMM, Transformer, MSTCN++) that are attempting the automated assessment of rehabilitation movement.

## Conclusion and Future Work

Through low-cost capture of clinic and home-based upper extremity therapy sessions, and the subsequent rating of these sessions by clinicians through our rating interface, we can produce data that increases the observability of the relation of movement quality to functionality. We can feed this data into our rehabilitation movement assessment HBM to quantify in a statistical manner the relations of movement quality to functionality. We can use the captured videos, clinician ratings and probabilities of our HBM to train computational algorithms for automated assessment of rehabilitation movement.

Automated assessment of rehabilitation movement during therapy would allow remotely supervised therapy at the home. We are creating an interactive therapy system (SARAH) [16] where a remote clinician can use the system to assign the sequence of tasks to be executed per training session at the home and receive automated summaries of performance. Clinicians can use automated summaries of therapy task performance at the home, along with quantitative identification of relations between movement changes and function for remote decision making in structuring therapy and remote feedback to the patient. We are planning to test the SARAH system in the home extensively thus generating more data for informing the HBM.

Automated assessment in the clinic can release more time for clinicians to focus on delivering therapy. The quantification of movement quality changes and their affect on functionality can help support the clinician decision processes for structuring therapy. In partnership with the Shirley Ryan Ability Lab (SRLab), we have adapted the SARAH system for use with automated assessment of the ARAT measure [23] in the clinic using four cameras. We have began the capture of over 100 patients performing the ARAT. The captures will generate standardized and invariable data of upper extremity rehabilitation movement while performing functional tasks. The captured data will further inform the HBM and test the transferability of our automated assessment approaches across the home and the clinic. Results from this work will be discussed in an upcoming publication.

Because our 15 training tasks (and their generalizable segment vocabulary) map well to ADLs, we expect that the relations established through our system between movement changes and task performance can reveal the relation between movement changes and overall daily life function. But the relation between the task layer of our hierarchy and the overall daily life functionality layer needs to be quantitatively developed and verified in a similar manner as the relation of the other layers. Some data for the ADL layer is already capturable through questionnaires as well as simple wearables (like activity monitoring applications on a smartphone). IMU based tracking of the affected limb, hand, and torso using methods being developed by our lab [34] and other research [8] can significantly increase observability of the daily activity. We are integrating unobtrusive IMUs units in the SARAH system [34]. The IMUs will be worn on the two wrists, index finger of the affected limb and on the waist throughout the day, including the training sessions at the home. The cross training of the IMU data with the video data and the expert ratings could allow us to identify particular distributions of IMU based kinematics that can be used to recognize particular segment blocks during ADLs. We could then adapt our decision tree algorithm to use this segment information to estimate types of tasks being performed.

We also plan to use the IMUs for localization(s). As certain types of tasks (i.e. reaching and grasping cups) have a high probability of being performed in certain spaces in the house (i.e. kitchen) we can use the localization as additional information for the computation of joint posterior probabilities of tasks and segments being performed during daily life activity. This would allow us to reconstruct the probabilities of all layers of the HBM during daily life (from task performance and location of performance to kinematics). We could then develop search algorithms that seek to continuously improve therapy impact for a patient by jointly maximizing the posterior probability of the HBMs calculated across therapy and ADL.

## Declarations

Ethics approval and consent to participate

IRB #97319 (Evaluating interactive systems for semi-automated assessment of movement functionality and movement quality of impaired subjects) was approved for impaired data capture at the Emory Hospital, Atlanta.

The following three IRBs were approved for unimpaired data capture at Virginia Tech.

- IRB #18-877 (Evaluating Interactive Systems for the Assessment of Movement Quality in Healthy Control Participants)
- IRB #19-920 (Understanding the activities and documentation needs of occupational and physical therapists)
- IRB #17-379 (Computational assessment of upper extremity movement) from video data

## Consent for publication

Not Applicable.

## Availability of data and materials

The datasets used and/or analysed during the current study are available from the corresponding author on reasonable request.

## Authors’ contributions

All the authors reviewed the manuscript before submission.

## Data Availability

All data produced in the present study are available upon reasonable request to the authors

## Acknowledgements

The authors would like to thank all the clinicians who participated in the rating sessions.

